# Clinical history shapes the predictive value of polygenic risk

**DOI:** 10.64898/2026.09.25.26364029

**Authors:** Dmitrii Usoltsev, Ivan Molotkov, Nikita Kolosov, Mary Pat Reeve, Alexander Ukhatov, FinnGen, Oxana Rotar, Anna Kostareva, Alexandra Konradi, Samuli Ripatti, Aarno Palotie, Mark J. Daly, Mykyta Artomov

## Abstract

For most diseases, whether PRS improves prediction beyond clinical data remains unknown, as existing evidence is concentrated in a handful of conditions with established risk models. Using 900,000 participants from UK Biobank and FinnGen, we developed and validated models for predicting 150 diseases and all-cause mortality. Genetic information provided the largest predictive gains in metabolic, cardiovascular, autoimmune, and neurological disease, but added little for genitourinary and respiratory conditions. All significant PRS-clinical risk interactions were negative, with predictive power gains from genetic data larger at lower clinical risk, suggesting that PRS may reveal susceptibility not yet apparent clinically. For example, in atherosclerotic cardiovascular disease, PRS expanded the clinically identified high-risk subgroup by 40%, and this reclassified subgroup had 4.9-fold higher incidence during the follow-up than those remaining low-risk. Our interpretable clinical-genetic models generalized across three biobanks and are publicly available through PrognosIQ.

## Introduction

Accurate prediction of disease onset can help identify individuals who may benefit from earlier screening, prevention, or clinical monitoring. Polygenic risk scores (PRS) provide a measure of inherited susceptibility across a broad range of common diseases [1–4]. In practice, genetic information is interpreted in the context of age, sex, medical history, comorbidities, medications, family history, and lab biomarkers [5–7]. Thus, a central question for clinical implementation of informative PRS testing is whether PRS adds meaningful information beyond an individual’s existing clinical profile.

Association of PRS with disease risk, however, is not the same as added clinical value. Clinical history and common laboratory biomarkers capture many observable risk factors and early disease-related changes, and some of these measures have substantial genetic architecture and are associated with disease outcomes [8]. As a result, PRS may add only modest information once clinical risk is already well characterized. For example, in coronary artery disease, adding PRS to established clinical risk scores has provided limited or modest improvements in predictive accuracy and risk stratification [9, 10]. At the same time, studies across multiple diseases have shown that high PRS is associated with higher lifetime risk and earlier disease onset, and that PRS can improve prediction beyond clinical risk scores for selected outcomes, including type 2 diabetes, breast cancer, and prostate cancer [11]. Disease-specific models, such as BOADICEA for breast cancer, further illustrate that integrating PRS with family history and rare pathogenic variants can improve individualized risk stratification [12]. Importantly, the added value of PRS may also vary across individuals. PRS performance differs substantially across genetic ancestry groups [13–16] and, even within a single ancestry group, varies by age, sex, and socioeconomic status [17].

These findings suggest that the predictive utility of PRS is likely to depend on broad disease context and the specific clinical model to which genetic information is added [18]. Existing studies of PRS integration into health outcome predictions have largely focused on a limited number of diseases with established clinical risk models, such as coronary artery disease or breast cancer [10–12]. In contrast, phenome-wide PRS studies evaluate genetic risk across many traits but typically benchmark PRS against simple covariates rather than rich clinical history [19]. Similarly, studies of PRS-by-clinical-factor interactions have generally focused on one disease area, or a limited set of exposures or outcomes [20–22]. Therefore, it remains unclear which diseases gain meaningful predictive information from PRS beyond clinical history, and for which subset of individuals this benefit is the greatest.

Here, we used clinical and genetic data from more than 900,000 individuals in UK Biobank [23] and FinnGen [24] to evaluate the value of PRS integration into clinical predictive models for the onset of 150 diseases and all-cause mortality. We first developed and externally validated transferable disease-onset models using clinical history, blood biomarkers, age, and sex, and benchmarked them against established clinical risk models where available. We then integrated disease-specific and multi-PRS models to quantify the incremental predictive value of genetics beyond clinical information. Finally, we performed a phenome-wide interaction analysis to test whether clinical phenotypes modify PRS effects and whether the predictive benefit of genetics differs across levels of clinical-history-based risk. Our results show that the added predictive power of PRS is not uniform across diseases or individuals, but is shaped by clinical background. In particular, PRS provided significant incremental benefit for a subset of diseases, with larger gains for metabolic, eye and ear disorders, autoimmune, neurological and cardiovascular outcomes, and often contributed most among individuals with lower clinical-history-based risk. To support translation, we implemented these models in PrognosIQ, a personalized disease-risk prediction platform integrating genetic risk, clinical history, and biomarkers.

## Results

### Clinical models for disease onset prediction show high transferability across biobanks

We modeled 151 outcomes (all-cause mortality + 150 diseases) in UK Biobank (**Sup. Tab. S1-S3, Sup. Fig. S1A; Fig. 1A**). Because observed associations between prior clinical diagnoses and future outcomes generally attenuated with time since diagnosis (**Sup. Tab. S4-S5; Sup. Fig. S1B**), we included time since diagnosis as potentially predictive features. The time-to-event was modeled using log-normal accelerated failure time models (AFT) [25], with features selected via a combination of forward step-wise selection and regularization [26] (**Sup. Tab. S6-S7, Sup. Fig. S1C**). For each outcome, we developed a null model including age and sex and a clinical model additionally incorporating selected clinical features, smoking, and BMI (**Methods. Clinical models development pipeline; Sup. Tab. S7; Fig. 1B**).

**Figure 1.**
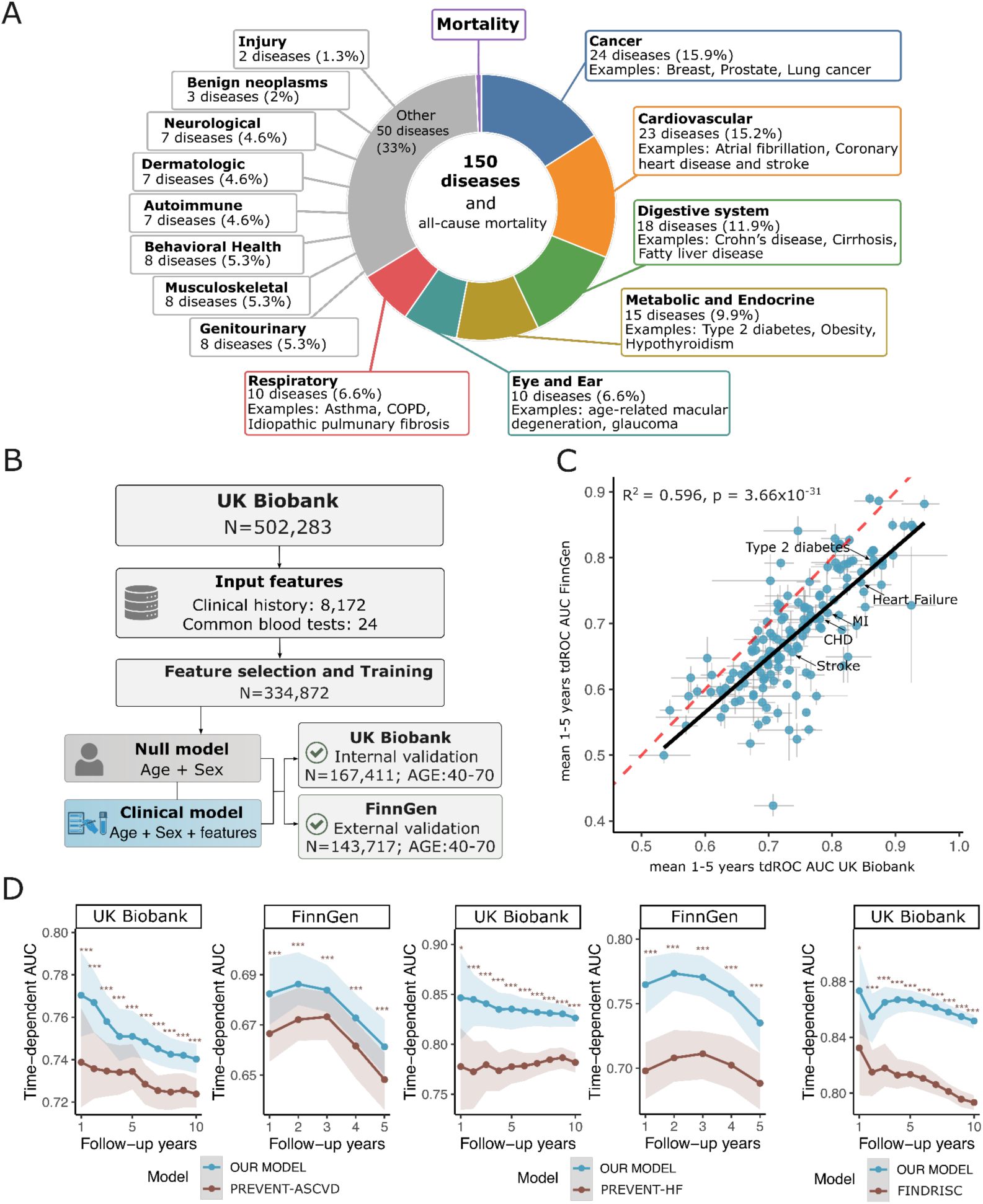
Development and validation of clinical disease-risk prediction models. **A) Distribution of the 150 selected diseases and all-cause mortality across disease categories.** Numbers and percentages indicate the number and proportion of modeled outcomes assigned to each category; representative diseases are shown for selected categories. **B) Workflow for clinical model development in UK Biobank and validation in FinnGen.** 8,172 Clinical-history features and 24 standard blood biomarkers were considered as candidate predictors. Models were trained in 334,872 UK Biobank participants aged 40-70 years and evaluated in an independent UK Biobank validation set (N = 167,411) and in FinnGen (N = 143,717). The null model included age and sex, whereas the clinical model additionally incorporated selected clinical-history and laboratory features. **C) Cross-biobank comparison of clinical model predictive performance across 150 diseases and all-cause mortality in UK Biobank and FinnGen.** Each point represents a disease evaluated in both cohorts; the x- and y-axes show the mean 1-5-year time-dependent receiver operating characteristic area under the curve (tdROC AUC) in UK Biobank and FinnGen, respectively. Error bars indicate 95% bootstrap confidence intervals estimated from 100 bootstrap resamples. The dashed diagonal line denotes equal performance between cohorts, and the solid line represents the linear regression fit across disease-specific performance estimates. The coefficient of determination and corresponding P value are shown in the panel (R² = 0.596, P = 3.66 × 10⁻³¹). **D) Benchmarking of the developed clinical models against established cardiovascular and type 2 diabetes risk-prediction models.** Time-dependent AUCs across follow-up horizons are compared with PREVENT-ASCVD for atherosclerotic cardiovascular disease outcomes, with PREVENT-HF for heart failure, and with FINDRISC for type 2 diabetes results. Shaded areas indicate 95% percentile bootstrap confidence intervals estimated from 100 bootstrap resamples. Statistical significance of differences between models was assessed using paired bootstrap comparisons based on the same bootstrap samples; *P < 0.05, **P < 0.01, and ***P < 0.001.

Clinical model performance was transferrable between UK Biobank and FinnGen (R² = 0.596, p = 3.66 × 10^-31^) (**Sup. Tab. S8)**, with 62 out of 151 models having time-dependent ROC AUC [27] (tdROC AUC) > 0.7 in both biobanks (**Fig. 1C**). Interestingly, the separate contributions of disease history and lab biomarkers were also transferable between the datasets (**Sup. Fig. S1D-E**). At the individual level, our clinical models showed stronger cross-biobank transferability than previously published embedding-based models, following the framework of Detrois et al. [28] (**Sup. Fig. S1F; Sup. Tab. S9**). Across biobanks, the resulting models outperformed established clinical risk scores-PREVENT-ASCVD [29], PREVENT-HF [29], and FINDRISC [30] - for cardiovascular disease, heart failure, and type 2 diabetes, respectively (**Fig. 1D, Sup. Fig. S1G**). Clinical model performance remained similar after excluding diagnoses obtained during the first year of follow-up, with strongly correlated estimates across outcomes (R² = 0.95, P = 2.74 × 10⁻⁹⁷), suggesting limited inflation due to diagnostic delay (**Methods. Estimation of models performance, Sup. Fig. S1H**).

Finally, we asked whether the clinical models could predict future disease among individuals without readily detectable signs of elevated risk, rather than simply identifying those already showing early clinical manifestations. Type 2 diabetes was selected as a representative example because individuals without prediabetes or major metabolic risk factors could be clearly defined and compared using the established FINDRISC score. We therefore evaluated model performance among UK Biobank participants with glucose levels <5.5 mmol/L, HbA1c <42 mmol/mol, BMI <30 kg/m^2^, and no use of diabetes-related medications. The model outperformed FINDRISC at longer follow-up times and was comparable in the shorter follow-up (**Methods. Estimation of models performance, Sup. Fig. S1I**).

### Genetic features improve prediction beyond clinical information, with heterogeneous gains across diseases

We evaluated two approaches for incorporating genetic information: a single disease-specific PRS and a multi-PRS model. Combining multiple polygenic scores, including scores for genetically correlated traits, has been shown to improve prediction accuracy [31]. For the single-PRS approach, we selected the PGS Catalog score with the highest ROC AUC in PGS Browser [3]; multi-PRS models were selected in FinnGen using forward stepwise Cox models adjusted for age, sex, and six genetic principal components (**Sup. Tab. S10**). Selected PRSs were then used in UK Biobank to construct genetic models with age and sex and clinical-genetic models additionally incorporating clinical features, smoking, and BMI. To minimize overlap bias, PRSs derived from FinnGen GWAS were excluded (**Methods. Genetic and clinical genetic predictive models; Sup. Tab. S11-S12; Fig. 2A**).

**Figure 2.**
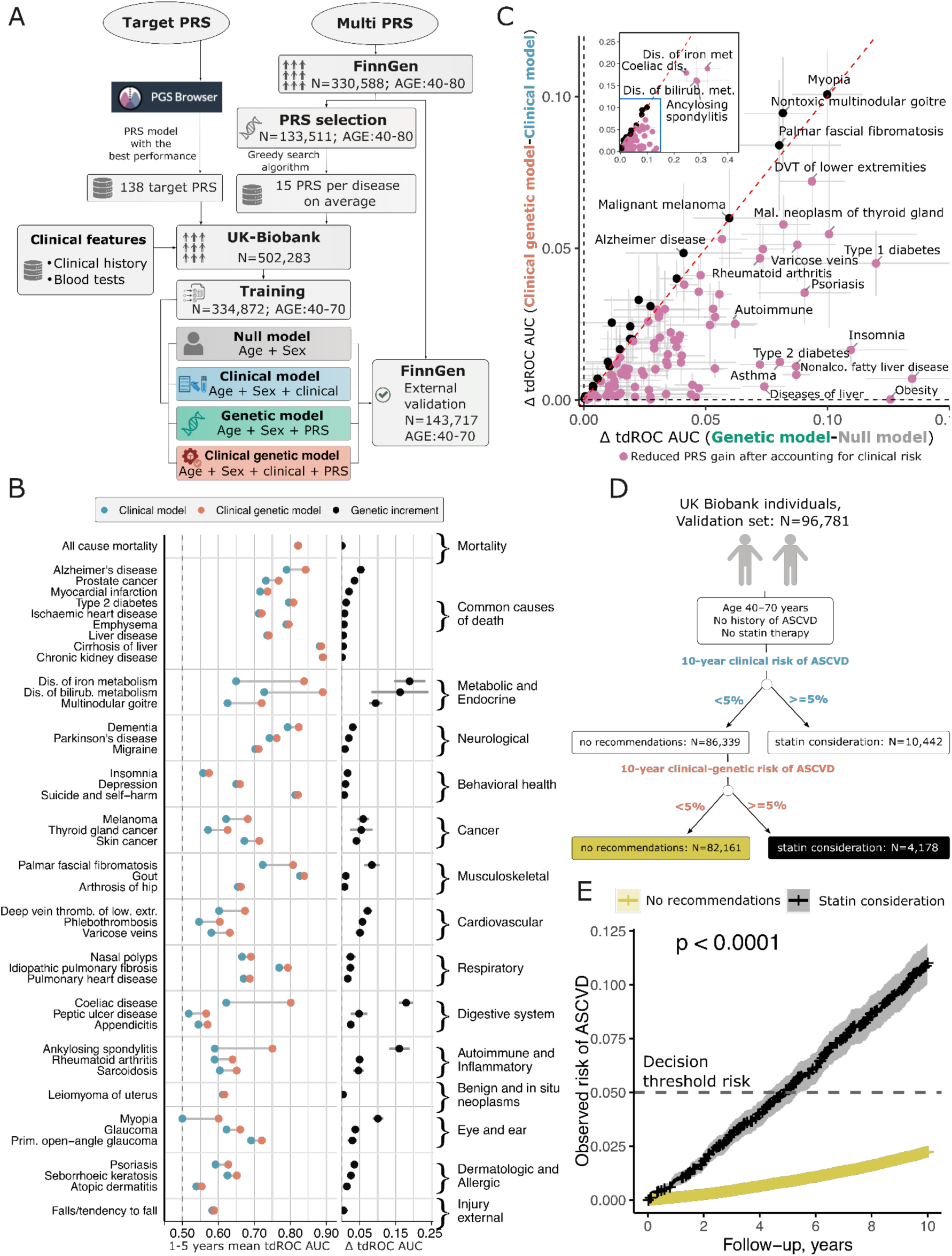
Development and evaluation of clinical-genetic models. **A) Overview of the model-development framework.** Disease-specific target polygenic risk scores (PRSs) were selected from the PGS Browser. Disease-specific combinations of PRSs were selected in FinnGen using a greedy search procedure, yielding an average of 15 PRSs per disease. Clinical predictors comprised disease-history variables and blood-test measurements from UK Biobank. Four model types were evaluated: null (age and sex), clinical (age, sex, and clinical predictors), genetic (age, sex, and PRSs), and clinical-genetic (age, sex, clinical predictors, and PRSs). Models were trained in UK Biobank (N = 334,872) and externally validated in FinnGen (N = 143,717), with both cohorts restricted to participants aged 40-70 years.**B) Comparison of clinical and clinical-genetic models in the FinnGen external validation cohort.** The left panel shows the mean 1-5-year tdROC AUC for the clinical and clinical-genetic models across representative diseases. The right panel shows the genetic increment, defined as the difference in tdROC AUC between the clinical-genetic and clinical models. Horizontal error bars represent 95% Wald confidence intervals. Diseases with a positive genetic increment and a FDR-adjusted Wald *P* value < 0.05 are shown, along with prespecified common causes of death. **C) Incremental predictive performance of adding PRS beyond clinical history.** The x-axis represents the increase in tdROC AUC of the genetic model relative to the null model, whereas the y-axis represents the increase in tdROC AUC from adding PRS to the clinical model (clinical-genetic versus clinical model). Each point represents a disease, and the horizontal and vertical error bars indicate the corresponding 95% Wald confidence intervals. Pink points denote diseases in which the incremental contribution of PRSs was reduced after accounting for clinical risk factors. The dashed diagonal indicates equal tdROC AUC increment in the two model comparisons; the inset magnifies the region containing diseases with larger performance gains. **D) Reclassification of 10-year ASCVD risk after incorporation of genetic information in the UK Biobank validation cohort.** Among 86,339 individuals classified as low risk (<5%) by the clinical model, 4,178 (4.8%) were reclassified upward to ≥5% by the clinical-genetic model, whereas 82,161 (95.2%) remained below the 5% decision threshold. **E) Kaplan-Meier estimates of cumulative ASCVD incidence among individuals initially classified as low risk (<5%) by the clinical model, stratified according to their classification after incorporation of genetic information.** Individuals reclassified upward to ≥5% had a substantially higher observed ASCVD risk than those who remained below the 5% threshold, reaching approximately 11.0% versus 2.2% cumulative incidence at 10 years (∼4.9-fold difference; P < 0.0001).

Clinical-genetic models outperformed clinical models on average in 15 of 16 disease groups, with urogenital diseases as the only exception (**Sup. Tab. S13-S14**). Although addition of genetic information generally improved predictive performance, the magnitude of improvement varied substantially across diseases. A significant positive increment attributable to genetic features was observed for 97 diseases. The largest mean increments were found for autoimmune diseases (N = 6, Δ tdROC AUC 0.054 ± 0.022), metabolic diseases (N = 11, 0.053 ± 0.020), gastrointestinal (N = 9, 0.036 ± 0.019), eye and ear disorders (N = 4, 0.045 ± 0.019), musculoskeletal (N = 4, 0.027 ± 0.019), cancer (N = 12, 0.026 ± 0.005), neurological diseases (N = 5, 0.028 ± 0.006), cardiovascular (N = 16, 0.027 ± 0.005), and common causes of death (N = 9, 0.016 ± 0.006) (**Sup. Fig. S2-3; Fig. 2B; Sup. Tab. S12-S16**).

Studies evaluating PRS predictive performance often measure improvement over age and sex alone, but these gains may not reflect the added value of genetics once clinical information is included. We therefore compared genetic predictive increments over null and clinical models across diseases (**Methods. Comparison of model performance, Fig. 2C; Sup. Tab. S17**). For 81.4% of diseases, the genetic increment was smaller in clinical-genetic models than in genetic models, although genetic predictive increments over null models and clinical models were moderately correlated across diseases (Spearman *ρ* = 0.65). Several phenotypes retained large PRS contributions despite incorporation of clinical information, most notably myopia (*Δ*AUC = 0.101), multinodular goiter (*Δ*AUC = 0.095), Dupuytren disease (*Δ*AUC = 0.084), and melanoma (*Δ*AUC = 0.060), with similar patterns across several cancers (**Sup. Tab. S17**). In contrast, examples where genetic gains were strongly attenuated in the presence of clinical history include obesity (0.126 to 0.0002), type 2 diabetes (0.087 to 0.011), and liver disease (0.074 to 0.0044), indicating substantial overlap between genetic and clinical predictive information (**Sup. Tab. S17**).

To examine whether genetic information could alter risk-based clinical decisions, we used ASCVD as a clinically actionable example. ASCVD prevention guidelines use the threshold of 5% for 10-year risk to consider lipid-lowering therapy [32]. We therefore used 5% as a guideline-informed reference threshold to evaluate whether adding genetic information identifies individuals whose estimated 10-year risk crosses a clinically relevant level. The analysis included 96,781 UK Biobank participants aged 40-70 years with no history of ASCVD and no prior statin therapy from the held-out validation set (**Fig. 2D**). Using the clinical model alone, 10,442 participants were identified at >=5% risk, corresponding to a guideline-relevant threshold for consideration of lipid-lowering therapy, while 86,339 participants had a predicted 10-year ASCVD risk below 5% and would therefore not have been identified for intervention. After incorporating genetic information, 4,178 of these low-risk individuals were reclassified to a predicted risk of ≥5%. Importantly, Kaplan-Meier analysis showed that individuals reclassified from <5% to ≥5% had a 4.9-fold higher cumulative incidence of ASCVD over the 10-year follow-up, than those who remained below the 5% threshold (P < 0.0001; **Fig. 2E, Sup. Fig. S4-S5**), indicating that genetic information identified a high-risk subgroup that would have been missed by the clinical model alone. We further assessed the robustness of this finding in FinnGen (**Sup. Fig. S6**), where individuals reclassified from <5% to ≥5% had a 1.9-fold higher cumulative ASCVD incidence by 6 years than those who remained below the 5% threshold (P < 0.0001).

### Polygenic risk contributes most to disease prediction among individuals with low clinical risk

To illustrate how polygenic risk varies with pre-existing clinical burden we used type 2 diabetes as an example. 418,391 FinnGen participants without diabetes at enrollment were considered. The T2D PRS showed significant interactions with baseline hypertension, dyslipidemia, and obesity, with consistently stronger effect of PRS on the T2D risk among individuals without these conditions (**Methods. Interaction study of Type 2 diabetes in FinnGen; Fig. 3A; Sup. Tab. S18**). Consistently, the hazard ratio per SD of PRS declined from 1.51 (95% CI, 1.48-1.53) in individuals without metabolic conditions to 1.28 (95% CI, 1.21-1.36) in those with at least two.

**Figure 3.**
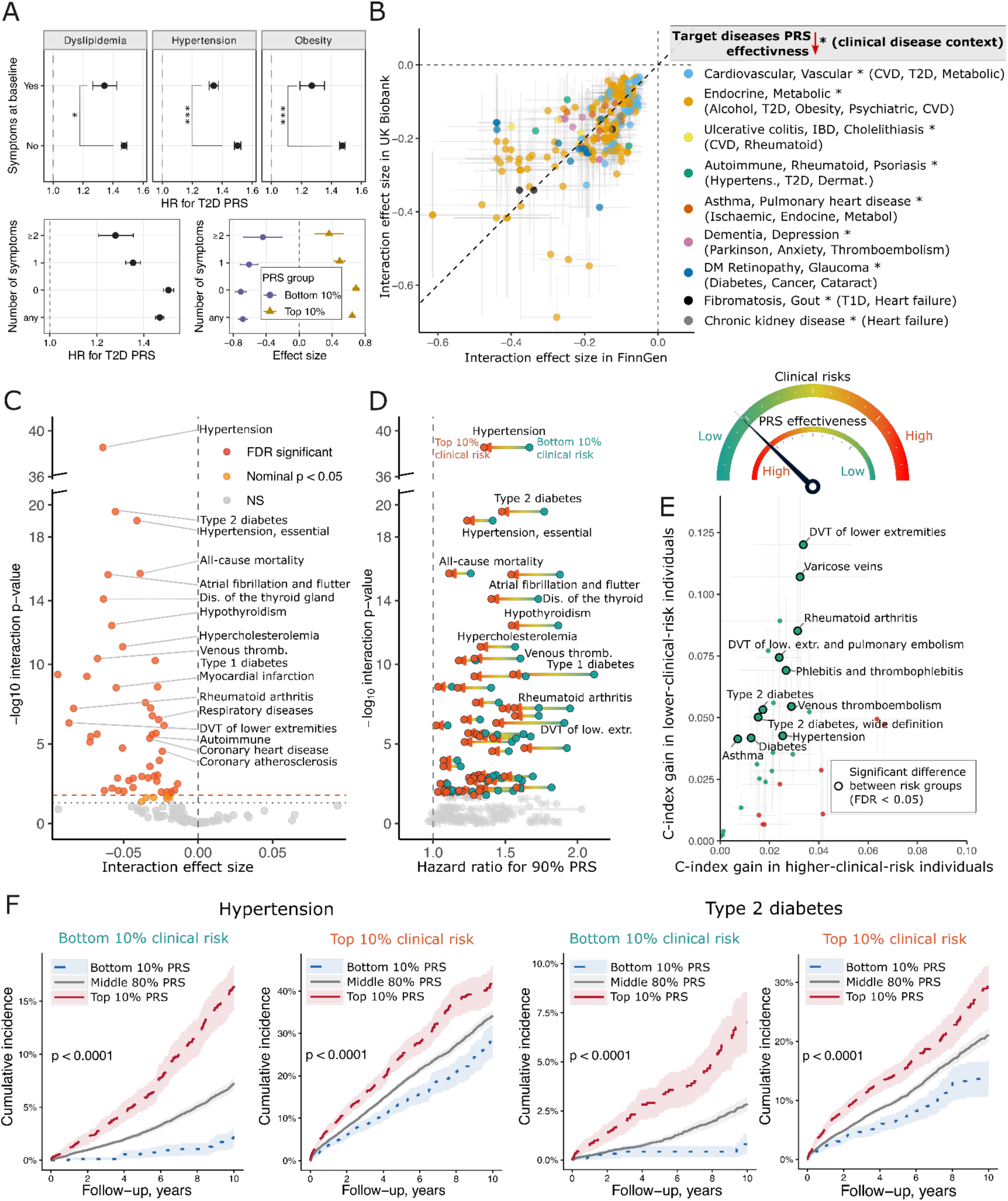
Clinical phenotypes modulate the effect of polygenic risk across diseases. **A) Modification of the association between the type 2 diabetes (T2D) PRS and disease risk by baseline clinical burden.** Hazard ratios (HRs) for the T2D PRS are shown according to the presence or absence of dyslipidemia, hypertension, and obesity, and according to the number of these risk factors. The lower-right panel illustrates differences in interaction effect estimates between individuals in the bottom and top 10% of the PRS distribution.**B) Identification of consistent interactions between clinical history and PRS in UK Biobank and FinnGen.** Visualisation of clinico-genetic interaction effects across biobanks. Each point represents a clinical phenotype × target-PRS interaction, with interaction effect estimates in FinnGen shown on the x-axis and corresponding estimates in UK Biobank on the y-axis. Horizontal and vertical error bars indicate 95% confidence intervals for the interaction effect estimates in FinnGen and UK Biobank, respectively. The dashed diagonal line indicates equal interaction effect sizes in the two cohorts.**C) Identification of interactions between clinical and genetic risk across 138 diseases and all-cause mortality.** The x-axis shows the estimated interaction effect and the y-axis shows -log10 of the nominal P value for the interaction term. Interactions passing FDR correction, nominally significant associations (P<0.05), and nonsignificant associations are indicated separately. Negative interaction effects indicate attenuation of the relative PRS effect with increasing clinical risk. Representative diseases with the strongest interaction signals are labeled. **D) Assessment of the modification of PRS effects by clinical risk.** For each disease, the PRS-associated hazard ratio was estimated at the 10th (low clinical risk) and 90th (high clinical risk) percentiles of the standardized clinical risk predictor. Arrows indicate the change in the PRS effect from low to high clinical risk, with right-to-left arrows representing attenuation of the PRS effect as clinical risk increases. The y-axis shows −log10 of the nominal P value for the PRS × clinical-risk interaction. Interaction P values were adjusted for multiple testing using the Benjamini-Hochberg procedure; colored arrows indicate interactions significant at FDR < 0.05. **E) Comparison of the gain in predictive performance achieved by adding genetic information to the clinical model across clinical-risk strata.** Individuals were divided into lower- and higher-clinical-risk groups using the median clinical risk as the cutoff. The x-axis shows the C-index gain of the clinical-genetic model over the clinical model among individuals with higher clinical risk, whereas the y-axis shows the corresponding gain among individuals with lower clinical risk. The dashed diagonal indicates equal genetic contributions in the two groups; points above and below the diagonal indicate greater incremental benefits of genetic information in the lower- and higher-clinical-risk groups, respectively. Large points with black outlines denote diseases with a significant difference in C-index gain between the two groups (FDR-adjusted Wald *P* < 0.05); smaller points denote nonsignificant differences.**F) Kaplan-Meier cumulative-incidence curves for hypertension and type 2 diabetes among FinnGen participants aged 40-70 years, stratified by clinical risk (bottom and top 10%) and PRS (bottom 10%, middle 80%, and top 10%).**

We next tested whether this attenuation generalized across diseases by evaluating 161,461 PRS-clinical-history interactions across 138 diseases in FinnGen (**Methods. Phenome-wide interaction study in FinnGen and UK Biobank, Sup. Tab. S19**). To avoid bias from GWAS and interaction sample overlap, FinnGen was used as the discovery cohort and PRSs derived using FinnGen samples were excluded; such overlap can substantially inflate type-I error in PGS interaction analyses [33]. This analysis identified 652 FDR-significant (FDR < 0.05) interactions, of which 587 were eligible for replication in UK Biobank and 277 were successfully replicated (**Sup. Tab. S20)**. Among the replicated interactions, only 11 had positive effect estimates, most involving pregnancy-related clinical codes whereas the remaining 266 - predominantly involving prior disease history - showed negative interactions between the clinical-history phenotype and the target PRS (**Sup. Fig. S7**), indicating attenuation of PRS effect in the presence of these phenotypes (**Fig. 3B**). Consistent with the expected effect on type-I error of sample overlap, using UK Biobank for interaction discovery yielded more than 11-fold as many significant interactions as the non-overlapping FinnGen analysis (**Methods. Phenome-wide interaction study in FinnGen and UK Biobank; Sup. Tab. S21**).

Because most replicated interactions indicated that adverse clinical-history phenotypes attenuated the effects of PRS, we next asked whether the same pattern was observed for overall clinical risk. We evaluated interactions between disease-specific clinical risk scores, derived from our clinical models, and the corresponding target PRS across 138 diseases and all-cause mortality (**Methods. Interaction between target disease PRS and predicted clinical risk in FinnGen)**. FDR-significant interactions were identified for 49 diseases and all-cause mortality, all of which had negative interaction effects. The most significant negative interactions were observed for hypertension (β = −0.064, p = 2.81 × 10-39), type 2 diabetes (β = −0.055, p = 2.60 × 10-20), all-cause mortality (β = −0.039, P = 1.99 × 10-16), atrial fibrillation and flutter (β = −0.060, p = 2.36 × 10-16), hypothyroidism (β = −0.058, p = 3.70 × 10-13), hypercholesterolemia (β = −0.051, p = 7.96 × 10-12), venous thromboembolism (β = −0.067, p = 4.37 × 10-11), and myocardial infarction (β = −0.055, p = 2.98 × 10-9) (**Fig. 3C; Sup. Tab. S22**). Importantly, sensitivity analyses excluding participants receiving statins, antihypertensive medications, or diabetes medications showed that the negative interactions remained consistent, indicating that they were not driven by baseline medication use (**Sup. Tab. S22, Sup. Fig. S8)**. The results also remained consistent after additionally accounting for PRS-by-age and PRS-by-sex interactions (**Sup. Tab. S22, Sup. Fig. S9**).

Consistent with these interactions, the estimated effect of a PRS at the 90th percentile relative to the mean was substantially larger at the 10th than at the 90th percentile of clinical risk - for example, the hazard ratio decreased from 1.67 to 1.35 for hypertension, 1.77 to 1.47 for type 2 diabetes, 1.88 to 1.54 for atrial fibrillation and flutter, and 1.60 to 1.29 for venous thromboembolism (**Fig. 3D; Sup. Tab. S22**).

To assess whether these interactions were reflected in differences in predictive performance, we compared the C-index gains from adding genetic information between individuals below and above the disease-specific median clinical risk score. Of the 49 diseases with significant interactions, 33 had sufficient cases and a significant genetic tdAUC increment for this analysis. Adding genetic information yielded greater C-index gains among individuals below the median clinical risk score for 25 of the 33 diseases. All 11 diseases with an FDR-significant between-group difference showed a greater predictive benefit from genetic information in the lower-clinical-risk group (**Methods. Comparison of the incremental predictive value of genetic information across clinical-risk strata. Fig. 3E; Sup. Tab. S23**). Consistent with this, Kaplan-Meier analyses for hypertension and type 2 diabetes showed that PRS separated disease trajectories even among individuals in the lowest decile of clinical risk, identifying substantial heterogeneity among people who would otherwise appear clinically similar (10-year cumulative incidence for T2D: 0.8% in the low-PRS group vs 7.0% in the high-PRS group; hypertension: 2.3% vs 16.4%), whereas the relative separation was smaller among individuals at high clinical risk (T2D: 14.0% vs 29.3%; hypertension: 28.0% vs 42.3%) (**Fig. 3F**). Overall, these findings highlight the potential of PRS to identify high-risk individuals before conventional clinical risk factors are observed.

To further test whether the predictive contribution of PRS diminishes with increasing clinical risk, we evaluated interactions between disease-specific PRS and a broad range of clinical, behavioral, dietary, and biomarker-related factors in UK Biobank across these 11 diseases. Among 1,411 FDR-significant interactions, factors associated with higher disease risk generally weakened the PRS effect, whereas protective factors tended to strengthen it. Accordingly, the main effects of these factors were inversely correlated with their interaction effects on PRS performance (Pearson R = -0.53, p < 1×10^-16^, **Sup. Tab. S24**).

### Translation of clinical genetic models into individualized disease prediction

The greater incremental value of genetic information among individuals with lower clinical risk suggests that PRS may help identify genetically susceptible individuals whose risk would not be apparent from conventional clinical assessment alone. However, the public access to externally validated models that integrate genetic susceptibility with detailed clinical history and biomarkers remains limited. To address this translational gap, we made the resulting models available through PrognosIQ, an individualized disease-risk prediction platform that combines population-adjusted PRS, medical history and blood biomarkers to estimate disease-specific absolute risk over time (**Methods. PrognosIQ web application for individualized risk prediction, Fig. 4A, Sup. Tab. S25**).

**Figure 4.**
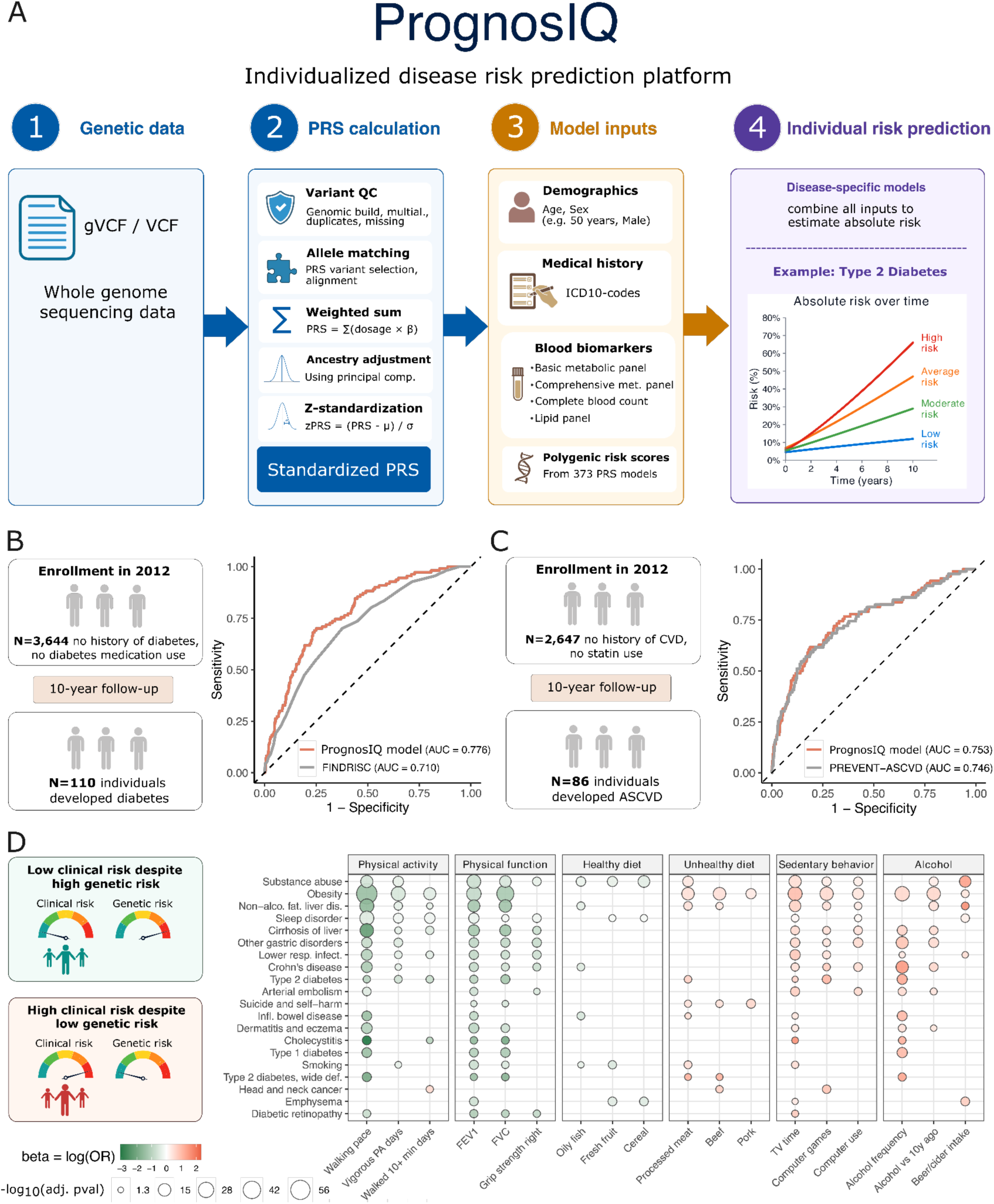
Translating integrated clinical and genetic risk models. **A) Overview of the PrognosIQ workflow.** Whole-genome sequencing data in gVCF/VCF format undergo variant quality control, allele matching, PRS calculation as a weighted sum of risk-allele dosages, ancestry adjustment using genetic principal components, and standardization. Standardized PRSs are integrated with medical history and blood biomarkers. Disease-specific models combine these inputs to estimate individualized absolute disease risk over time. **B) Evaluation of PrognosIQ for prediction of 10-year type 2 diabetes risk.** The analysis included 3,644 individuals without a history of diabetes and without use of diabetes medication at enrollment in 2012; 110 individuals developed diabetes during 10 years of follow-up. Receiver operating characteristic (ROC) curves compare PrognosIQ (AUC=0.776) with FINDRISC (AUC=0.710). **C) Evaluation of PrognosIQ for prediction of 10-year atherosclerotic cardiovascular disease (ASCVD) risk.** The analysis included 2,647 individuals without a history of cardiovascular disease and without statin use at enrollment; 86 individuals developed ASCVD during 10 years of follow-up. ROC curves compare PrognosIQ (AUC=0.753) with PREVENT-ASCVD (AUC=0.746). **D) Lifestyle and clinical characteristics of individuals with discordant clinical and genetic risk profiles**. Associations are compared between individuals with low clinical risk despite high genetic risk and those with high clinical risk despite low genetic risk. Rows represent clinical phenotypes and columns represent lifestyle characteristics grouped into physical activity, physical function, healthy diet, unhealthy diet, sedentary behavior, and alcohol consumption. Circle color represents the direction and magnitude of the association coefficient and circle size is proportional to −log10 of the adjusted P value.

We externally evaluated PrognosIQ in an independent prospective cohort that differs from the training datasets in healthcare setting, geographic location, and ancestry composition and was previously described by Usoltsev et al. [34]. Among 3,644 participants who were free of type 2 diabetes at baseline in 2012, PrognosIQ achieved an ROC AUC of 0.776 for predicting incident type 2 diabetes, compared with 0.710 for FINDRISC (**Fig. 4B**). We also evaluated ASCVD prediction in 2,647 participants with no history of cardiovascular disease and no statin use at baseline, where PrognosIQ achieved an ROC AUC of 0.753, compared with 0.746 for the PREVENT-ASCVD equation. Notably, all variables required to calculate FINDRISC and PREVENT-ASCVD were available, whereas PrognosIQ predictions were generated using only the limited subset of clinical data available in this cohort (**Methods. External validation of PrognosIQ, Fig. 4C**).

Finally, we asked whether potentially modifiable factors distinguish individuals whose clinical risk differs from their inherited susceptibility **(Methods. Phenotype analysis of discordant clinical and genetic risk groups)**. Across multiple diseases, lower clinical risk among genetically susceptible individuals was associated with greater physical activity and better physical function, including a faster walking pace, more frequent vigorous activity and stronger grip, as well as indicators of a healthier diet (**Fig. 4D, Sup. Tab. S26**). Conversely, high clinical risk despite low genetic susceptibility was associated with unhealthy dietary patterns, sedentary behavior, and alcohol-related characteristics. These associations were observed across a broad range of outcomes, including obesity, type 2 diabetes, cardiovascular and respiratory diseases, and substance-related disorders. Together, these findings highlight lifestyle and physical function as modifiable factors that may contribute to attenuation of elevated inherited disease risks.

## Discussion

Our findings address two questions that are central to the clinical use of PRS: for which diseases does genetic information improve prediction beyond routinely available clinical data, and in which patients is that improvement greatest? Using genetic and longitudinal clinical data from more than 900,000 participants in UK Biobank and FinnGen, we developed predictive models for 150 diseases and all-cause mortality and systematically evaluated how genetic information contributes to prediction across diseases and clinical contexts. The resulting clinical-genetic models are publicly available through PrognosIQ, a platform integrating clinical history, biomarkers, and genetic information into individualized disease-risk estimates. Across outcomes, the contribution of PRS varied from substantial to nearly absent and frequently depended on an individual’s clinical background. Our results show that the predictive utility of PRS cannot be inferred from its association with disease alone and should instead be evaluated within the clinical model and patient population in which it would be used. This provides a basis for prioritizing PRS implementation in diseases and patient groups where genetic information meaningfully improves prediction beyond routinely available clinical information.

PRS produced the clearest improvements in prediction for cardiovascular, autoimmune, metabolic, and cancer outcomes, including several of the best-studied examples of polygenic risk prediction, such as atrial fibrillation, breast and prostate cancer [11]. However, for many diseases, including liver disease and obesity, PRS added little predictive utility beyond clinical history, despite strong univariate associations with disease risk. Prior studies have reported a similar distinction between genetic association and improvement over clinical models; for example, adding a coronary artery disease PRS to pooled cohort equations produced only a modest increase in discrimination and changed estimated risk minimally for most participants [9]. Asthma was a notable example in our analysis, with a significant PRS association but limited improvement in prediction. This may partly reflect the clinical and biological heterogeneity of asthma, as childhood- and adult-onset disease have partly distinct genetic architectures and different relationships with obesity, smoking, and other exposures [35]. More generally, small gains may occur when clinical diagnoses, biomarkers, and medication history already reflect downstream consequences of genetic susceptibility.

The interaction analyses further showed that the contribution of PRS varied according to prior disease history, socioeconomic characteristics, and overall risk estimated from clinical history. Genetic ancestry is already well established as a major source of heterogeneity in PRS performance [13–16]; here, we focused on an additional and less systematically characterized source of variation - differences in clinical background among individuals of similar ancestry. Our findings extend earlier studies showing that PRS predictive performance varies with age, sex, and socioeconomic background [17], and that associations between polygenic risk and disease can vary across environmental and clinical contexts [18, 20, 22]. Our study’s cross-biobank design was particularly important for the interaction analyses, because an overlap between PRS discovery GWAS and interaction samples can inflate type-I error in interaction analyses [33]. Consistent with this concern, using UK Biobank for interaction discovery yielded more than 11-fold as many significant interactions as the non-overlapping FinnGen analysis, despite the higher disease prevalence and discovery power previously reported for FinnGen relative to UK Biobank for common disease endpoints [24]. The cross-biobank consistency of the main findings therefore argues against the observed patterns being specific to a single cohort or healthcare system.

The most consistent pattern was that PRS improved prediction more among individuals classified as low risk by the clinical models. One possible explanation is partial redundancy between inherited susceptibility and its clinically observable downstream manifestations: as disease-related diagnoses and biomarkers accumulate, a greater fraction of genetically correlated risk becomes represented in the clinical profile. This pattern suggests a potential role for PRS in identifying susceptibility before elevated risk becomes clinically apparent. Because inherited risk is available before many diagnoses, treatments, and biomarker abnormalities emerge, PRS may be particularly informative when the clinical record contains few indicators of future disease. Previous studies have shown that PRS can identify individuals with elevated lifetime risk or earlier disease onset who may be missed by conventional risk factors [11, 36, 37]; our results extend this observation across a broad range of outcomes. Clinical implementation, however, will require disease-specific evaluation of calibration, intervention thresholds, available preventive measures, and potential harms of additional screening.

This study has several limitations. First, UK Biobank and FinnGen predominantly include individuals of European ancestry and represent specific healthcare systems, which may limit the generalizability of both the clinical models and PRS effects to other populations. Second, clinical history was derived from available diagnoses, medications, biomarkers, and other biobank records and may incompletely capture risk factors such as family history, lifestyle, and detailed physiologic measurements. These records may also reflect differences in healthcare utilization, and some features recorded shortly before disease onset could represent early manifestations of disease rather than independent predictors. Third, limited improvement for a given disease can be due to poor performance of existing PRS models and may not necessarily indicate that inherited susceptibility is clinically uninformative. Finally, although PrognosIQ enables implementation of the resulting models, prospective studies are needed to assess calibration in clinical populations and determine whether PRS-informed screening or prevention improves patient outcomes.

Overall, these findings support a selective and clinically contextualized approach to utilizing PRS in clinical outcome prediction. PRS added meaningful predictive information for a defined subset of diseases, provided little improvement for many others, and often had its greatest value among individuals with limited evidence of risk in their clinical history. The broad disease coverage, replication across two biobanks, and systematic evaluation of interactions together provide a framework for identifying both the outcomes and patient groups in which genetic information is most likely to improve risk assessment.

## Methods

### Clinical models development pipeline

#### a) Harmonization of UK Biobank and FinnGen clinical history phenotype definitions

UK Biobank data freeze from December 25, 2023, was accessed through the UKB-RAP platform and contained information on 502,283 individuals. Individual-level disease information was obtained from hospital inpatient records (fields 41270 and 41280), UK Biobank-derived first-occurrence fields (fields 130000-132604), which integrate information from hospital admissions, primary care, death records, and self-reported conditions, and cancer registry data (fields 40006 and 40008). These sources were combined to derive, for each participant, recorded ICD-10 diagnoses and the earliest available age at which each condition was documented.

UK Biobank clinical-history phenotypes were then harmonized with FinnGen clinical endpoints using ICD-10 mappings derived from the FinnGen DF11 endpoint definitions (**Sup. Materials. Harmonization of UK Biobank clinical-history phenotypes with FinnGen endpoint definitions, Sup. Tab. S1)**.

All longitudinal registry data were censored on 19 December 2022. Dates of birth were approximated using the year and month of birth recorded in fields 34 and 52, with the day set to the 15th of the corresponding month. Baseline age was calculated as the interval between the estimated date of birth and the date of the baseline assessment (field p53_i0). Age at event or censoring was calculated as the interval between the estimated date of birth and the earliest of the target-event date, censoring date, or date of death (field 40000_i0). All time intervals were calculated in R using the difftime function, expressed in weeks, and divided by 52.25 to convert them to years. Follow-up time was calculated as age at event or censoring minus baseline age.

Laboratory measurements were also harmonized between UK Biobank and FinnGen **(Sup. Materials. Harmonization of laboratory measurements between FinnGen and UK Biobank).**

#### b) Disease selection

A total of 3,791 clinical endpoints in UK Biobank were matched with respective ICD-10-based FinnGen phenotype definitions to ensure harmonization across the two biobanks. For disease selection, each endpoint was required to include more than 500 incident cases occurring after baseline assessment age. For the future comparisons of predictive performance upon inclusion of genetic data, we also required that an established polygenic risk score model was available in PGS Browser [3] for the disease of interest. 138 diseases passed these inclusion criteria. When multiple PRS were available for the same disease, the best-performing score was selected based on the lifelong risk prediction quality reported in PGS Browser [3]. The final outcome set comprised all-cause mortality and 150 diseases, including 11 additional endpoints reported by the US Centers for Disease Control and Prevention (CDC) as major contributors to mortality [38] and macular degeneration, which was included as an additional clinically relevant endpoint.

#### c) Cox model with time varying coefficients and time-dependent features

To characterize the individual association of each clinical-history phenotype with subsequent disease risk, we first performed a series of univariable Cox analyses [39] for each of the selected disease outcomes. Each clinical-history phenotype was modeled as a time-dependent exposure. For each disease outcome, follow-up extended from baseline age until the age at disease onset or censoring. Only phenotypes that occurred after baseline and before disease onset were considered time-dependent exposures.

Follow-up was divided into intervals according to time since onset of the clinical-history phenotype: 0-1, 1-3, 3-5, 5-7, 7-10, and >10 years. For individuals who did not develop the clinical-history phenotype before disease onset or censoring, all time-dependent exposure indicators were set to zero. For individuals who developed the phenotype during follow-up, the period before its onset was coded as unexposed, whereas subsequent follow-up was assigned to the corresponding time-since-onset interval.

Cox proportional hazards models were fitted using the Surv(tstart, tstop, event) formulation implemented in the R package survival (v3.4-0) [40]. Separate indicator variables were included for each time-since-onset interval, allowing the hazard ratio associated with each clinical-history phenotype to vary according to the time elapsed since phenotype onset. Models were adjusted for sex, except for sex-specific disease outcomes. For these outcomes, the analysis was restricted to participants of the relevant sex, and sex was omitted from the model. In UK Biobank, the analysis was performed separately for participants aged 40-50, 50-60, and 60-70 years. The analysis was independently replicated in a randomly selected subset of 157,879 FinnGen participants, stratified into the 40-50, 50-60, 60-70, and 70-80-year age groups (**Sup. Tab. S4-S5, Sup. materials. Analysis of time-dependent features**).

#### d) Clinical feature selection

First, we conducted univariate analyses in which each phenotype was used separately to predict the target outcome using a Cox model with time-varying coefficients and time-dependent features using R package survival (v3.4-0). Predictors that were not associated with the outcome in any time-since-onset interval (P > 0.05 in all intervals) were excluded. For pairs of highly correlated phenotypes (Pearson’s r > 0.8), the predictor with the higher univariate C-index was retained.

The next stage of feature selection was performed separately for each target disease and baseline age group (40-50, 50-60, and 60-70 years) using three complementary approaches: a baseline Cox proportional hazards model, a Cox model with time-dependent covariates, and logistic regression. Both Cox models were fitted in R using the survival package (v3.4-0). All approaches used forward selection. The baseline Cox and logistic regression models started from a model containing age and sex, whereas the time-dependent Cox model started from a model containing sex only. In the baseline Cox model, clinical-history phenotypes were included as fixed baseline covariates and coded as absent if their onset occurred after baseline. In the time-dependent Cox model, follow-up was represented in start-stop format, with exposure status updated according to phenotype onset and time since onset. In logistic regression, the target disease was modeled as a binary outcome, and clinical-history phenotypes occurring after disease onset were coded as absent to minimize temporal leakage. At each step, the predictor producing the largest improvement was added, provided that it increased the C-index by at least 0.001 in either Cox model or McFadden’s pseudo-R² [41] by at least 0.001 in logistic regression. Selection stopped when no remaining predictor met the corresponding threshold.

Features selected across the three baseline-age groups were combined into three nested candidate feature pools: phenotypes selected by the baseline Cox models alone; phenotypes selected by either the baseline or time-dependent Cox models; and phenotypes selected by any of the three feature-selection approaches. For each feature pool, we fitted a log-logistic accelerated failure time model with elastic-net regularization in Python 3.11 using lifelines v0.29.0 [42]. The relative contribution of L1 and L2 regularization was evaluated using L1 ratios of 0, 0.25, 0.5, 0.75, and 1.0. For each L1 ratio, the overall penalty strength was optimized within the interval from 0 to 0.01 using bounded scalar optimization implemented in Python 3.11 SciPy (v1.15.2) (scipy.optimize.minimize_scalar) [43]. Candidate models were fitted in the training set, and hyperparameters were selected by maximizing C-index in the held-out test set. In addition to the optimizer-derived value, penalty strengths of 0, 10⁻⁶, 10⁻⁵, 10⁻⁴, 10⁻³, and 10⁻² were evaluated explicitly. The combination yielding the highest C-index was selected. The model was subsequently refitted in the training set using the selected hyperparameters, and predictors with model-based P values <0.05 were retained. Finally, the model was refitted using the selected predictors and hyperparameters in the combined training and test sets.

Laboratory predictors were selected from commonly used biomarker panels, including the complete blood count, basic metabolic, comprehensive metabolic, and lipid panels (**Sup. Tab. S6**). For each target disease and baseline age group, analyses were restricted to participants in the training set who were within the corresponding age range at baseline and whose target-disease event, if present, occurred after baseline. Analyses of sex-specific outcomes were further restricted to participants of the relevant sex.

Within each age group, laboratory features were selected using forward stepwise selection. Starting from a baseline Cox model containing only age and sex as covariates, candidate laboratory measurements were evaluated one at a time, and the feature producing the largest increase in the C-index was added. A feature was retained only if it increased the C-index by at least 0.001; otherwise, selection was stopped. This procedure yielded age-group-specific sets of laboratory features.

Finally, laboratory features selected across age groups were combined and used to fit elastic-net-regularized log-logistic accelerated failure time models in Python 3.11 using lifelines v0.29.0. Hyperparameters were optimized in the test set, and features with model-based P values <0.05 were retained. The final AFT model was then refitted on the combined training and test sets using the selected features and optimized hyperparameter configuration.

#### e) Development of Null and Full Clinical Models

The null model was specified as a log-normal accelerated failure time (AFT) model including only age and sex. The full clinical model used the same log-normal AFT framework and additionally incorporated selected clinical-history phenotypes, laboratory measurements, body mass index, and smoking status, where available. All models were fitted in Python 3.11 using lifelines v0.29.0. After model specification and evaluation, the final model was refitted using the combined training and test sets.

### Estimation of models performance

#### a) Calculation of time-dependent ROC AUC

Model performance was evaluated separately for each target disease in held-out validation cohorts. In UK Biobank, the clinical and null models were evaluated in 167,411 participants. In FinnGen, the null, clinical, genetic, and clinical-genetic models were evaluated in 143,717 participants (**Fig. 2A)**. Individuals who developed the target disease before or at baseline were excluded.

Discrimination was quantified at annual prediction horizons using time-dependent ROC AUC (tdROC AUC) calculated with the timeROC R package (v0.4) [44]. Performance was evaluated only at horizons for which both cases and controls were available. Annual tdROC AUC values were calculated at prediction horizons from 1 through 5 years. To assess potential bias from diagnostic delay, we additionally evaluated model performance in FinnGen after excluding participants diagnosed with the target disease within the first year after baseline. In this sensitivity analysis, performance was summarized as mean tdROC AUC across prediction horizons of 2-5 years.

Uncertainty in performance estimates was assessed using nonparametric bootstrap resampling. Within each disease and prediction horizon, participants were sampled with replacement. The same bootstrap samples were applied to all models, preserving the paired structure of model comparisons. Point estimates were calculated in the original validation sample, and bootstrap distributions were used to estimate mean performance and percentile-based confidence intervals.

#### b) Transferability of individual-level predicted risk of clinical-history models between UK Biobank and FinnGen

To assess cross-cohort model transferability, the clinical model-development pipeline used in UK Biobank was independently repeated in FinnGen. The FinnGen cohort was divided into training (N = 157,879), testing (N = 157,903), and validation (N = 157,902) subsets, and disease-specific accelerated failure-time models were developed using baseline age, sex, and selected clinical-history phenotypes, as described above (**Methods. Clinical feature selection**). For each disease, linear predictors (the weighted sum of the model predictors for each individual with weights corresponding to the estimated effect sizes) from the FinnGen-trained model and the corresponding UK Biobank-trained model were generated for the same participants in the FinnGen validation set. Analyses were restricted to individuals aged 40-70 years at baseline whose event or censoring age was greater than their baseline age.

To remove the direct contributions of baseline age and sex, each linear predictor was subsequently adjusted by subtracting the model-specific terms using coefficients estimated in the corresponding fitted model (*β_a_*_g*e*_ × *baseline age* + *β_sex_* × *sex*). The adjusted linear predictors were then multiplied by −1 so that higher values corresponded to higher predicted disease risk.

Pearson correlation coefficients, two-sided P values, and 95% confidence intervals were then calculated between the adjusted linear predictors.

As an individual-level measure of model transferability, C-index was calculated separately for the adjusted FinnGen-trained and UK Biobank-trained linear predictors in the FinnGen validation set, using time from baseline to the event or censoring. Linear predictors were standardized before C-index estimation. This analysis was designed to provide an individual-level risk transferability assessment analogous to that described by Detrois et al. [28].

#### c) Calculation of the PREVENT score in UK Biobank and FinnGen

Ten-year risks of atherosclerotic cardiovascular disease (ASCVD) and heart failure were calculated using the base American Heart Association PREVENT equations implemented in the PooledCohort R package (v0.0.2) [45] (equation_version = "Khan_2023"). The models included age, sex, smoking status, total and HDL cholesterol, systolic blood pressure, antihypertensive and statin use, diabetes, body mass index, and estimated glomerular filtration rate (eGFR). Variables were harmonized across UK Biobank and FinnGen and converted to the units required by PREVENT. eGFR was calculated using the 2021 CKD-EPI creatinine equation [46]. Analyses were limited to participants within the supported predictor ranges. tdROC AUC was evaluated in held-out validation sets after excluding individuals with prevalent ASCVD at baseline. The 10-year PREVENT estimate was used as a fixed risk score across follow-up horizons.

For comparison with PREVENT-ASCVD, ASCVD risk was derived from three disease-specific prediction models: myocardial infarction, major coronary heart disease events, and stroke. For each participant and prediction horizon, the maximum predicted risk across the three models was used as the composite ASCVD risk estimate.

#### d) Calculation of the FINDRISC in UK Biobank

The Finnish Diabetes Risk Score (FINDRISC) was reconstructed using UK biobank measurements and questionnaire data obtained during the first visit. The score included eight components and ranged from 0 to 26 points. Age contributed 0 points for participants younger than 45 years, 2 points for those aged 45-54 years, 3 points for those aged 55-64 years, and 4 points for those aged over 64 years. Body mass index (field 21001_i0) contributed 0 points for values below 25 kg/m², 1 point for values of 25 to <30 kg/m², and 3 points for values ≥30 kg/m².

Waist circumference (field 48_i0) was scored using sex-specific thresholds. Men received 3 points for a waist circumference of 94 to <102 cm and 4 points for a waist circumference ≥102 cm; the corresponding thresholds for women were 80 to <88 cm and ≥88 cm. Participants reporting less than 30 minutes of combined moderate (field p894_i0) and vigorous physical activity (field 914_i0) received 2 points. Participants reporting no daily consumption of fresh fruit (field 1309_i0), dried fruit (field p1319_i0), raw vegetables (field p1289_i0), or cooked vegetables (field 1299_i0) received 1 point.

The use of antihypertensive medication (field 6177_i0) at baseline contributed 2 points. A history of elevated blood glucose contributed 5 points and was defined as baseline serum glucose (field 30740_i0) concentration ≥6.1 mmol/L. A first-degree family history of diabetes, defined as diabetes reported in a biological mother (field 20110_i0), father (field 20107_i0), or sibling (field 20111_i0), contributed 5 points. Information on second-degree relatives was not available in UK Biobank.

Additionally, we marked individuals who reported using diabetes-related medications in UK Biobank (field 20003_i0). These medications were identified using the following Anatomical Therapeutic Chemical (ATC) codes: A10BA, A10BB, A10BC, A10BD, A10BF, A10BG, A10BH, A10BJ, A10BX, A10AD30, A10AB01, A10AB04-A10AB06, A10AE01, A10AE04-A10AE06, and A10AE30.

The analysis was restricted to participants without type 2 diabetes before baseline and with complete information for all FINDRISC components. tdROC AUC was evaluated in held-out validation. The FINDRISC estimate was used as a fixed risk score across follow-up horizons.

### Genetic and clinical genetic predictive models

#### a) Genetic feature selection

A total of 2,692 unique PRS models developed without the use of FinnGen data were selected from the PGS Browser. PRS scores for FinnGen participants were obtained from the PGS Browser. To derive multi-PRS feature sets, genetic feature selection was performed separately for each disease and baseline-age stratum (40-50, 50-60, 60-70, and 70-80 years) in the FinnGen cohort, comprising 133,511 participants who were not included in the validation cohort. Analyses were restricted to individuals who had positive follow-up after baseline. For sex-specific diseases, analyses were further restricted to participants of the relevant sex.

In the first stage, all candidate PRSs underwent univariate association screening using Cox proportional hazards models adjusted for baseline age, sex, and the first six genetic principal components using R package survival (v3.4-0). PRSs that passed the Bonferroni-corrected significance threshold of P<0.05/2,692 and were available in the validation cohort were retained as candidate features.

Candidate PRSs were then selected using a forward stepwise selection procedure based on improvement of C-index in a Cox model that included baseline age, sex, and the first six genetic principal components as covariates. At each iteration, each remaining candidate PRS was added individually to the baseline covariates and any PRSs selected in previous iterations. The model producing the largest increase in the C-index relative to the current model was retained, and the corresponding PRS was added to the selected feature set. Selection continued until none of the remaining PRSs improved the C-index by at least 0.001. The final PRS feature set and the C-index obtained at each selection step were recorded separately for each disease and age stratum.

#### b) Development of genetic and clinical genetic predictive models

PRS features selected across baseline age strata in FinnGen were combined and used as candidate predictors to train genetic accelerated failure time (AFT) models with elastic-net regularization in Python 3.11 using the lifelines package (v0.29.0). Model hyperparameters were optimized using the UK Biobank test set, after which the final genetic models were refitted using the combined training and test datasets. PRS features retained in each final genetic model were subsequently combined with predictors from the corresponding clinical model to construct the final clinical-genetic AFT model.

For UK Biobank, PRS scoring files were downloaded and harmonized using PGS Catalog utilities [47, 48], and individual-level PRSs were calculated from imputed genotype data using PLINK 2.0 [49]. PRSs were adjusted for population structure and standardized before inclusion in predictive models (**Sup. materials. Calculation of PRS in UK Biobank**). FinnGen PRSs were adjusted for population structure before model estimation and downstream analyses, as described in the PGS Browser [3].

### Comparison of model performance

Predictive performance was compared across the null, clinical, genetic, and clinical-genetic models using time-dependent ROC AUC (tdROC AUC). For each endpoint, all four models were evaluated using identical bootstrap resamples, preserving the paired structure of model comparisons. For analyses spanning multiple prediction horizons, tdROC AUC values were averaged across the prespecified horizons within each bootstrap iteration before calculating differences between models.

Performance differences were estimated in the original validation sample, and their standard errors were calculated as the standard deviation of the corresponding paired bootstrap differences. Statistical significance was assessed using two-sided Wald tests, with the test statistic defined as the estimated difference divided by its bootstrap-derived standard error. The corresponding 95% Wald confidence intervals were constructed as the estimated difference ± 1.96 times this standard error. Reference models with tdROC AUC values below 0.5 were set to 0.5.

### Interaction study of type 2 diabetes in FinnGen

Analyses were restricted to FinnGen participants who were free of type 2 diabetes (T2D) at baseline and had positive follow-up. Follow-up time was calculated as the difference between age at T2D onset or censoring and baseline age. The population-structure-adjusted T2D PRS (PGSID: PGS002354 [50]) was standardized to a mean of 0 and a standard deviation of 1.

We examined whether the association between the T2D PRS and incident T2D differed depending on three baseline cardiometabolic conditions: hypertension, dyslipidemia, and obesity. Conditions with an onset after baseline were coded as absent. For each condition, a Cox model including the T2D PRS, condition status, their interaction, baseline age, and sex was fitted. The significance of effect modification was assessed using the p-value for the PRS-by-condition interaction term. To facilitate interpretation, separate Cox models adjusted for baseline age and sex were also fitted among participants with and without each condition, and hazard ratios for T2D per 1-standard-deviation increase in the PRS were estimated.

To assess the combined influence of baseline cardiometabolic burden, participants were grouped according to the number of these conditions present at baseline: 0, 1, or ≥2. Cox models adjusted for baseline age and sex were fitted overall and separately within each burden group to estimate the hazard ratio associated with a 1-standard-deviation increase in the T2D PRS. In an additional analysis, participants were classified using the 10th and 90th percentiles of the PRS distribution as the bottom 10%, middle 80%, and top 10% genetic risk groups. Within each cardiometabolic-burden stratum, Cox models adjusted for baseline age and sex were used to compare the bottom and top PRS groups with the middle 80% reference group. Hazard ratios and 95% confidence intervals were calculated from the model coefficients and their standard errors. All Cox models were fitted in R using the survival package (v3.4-0).

### Calculation of clinical risk on linear scale for interaction studies

For each disease, we fitted parametric Accelerated Failure Time (AFT) models with log-logistic and log-normal distributions, regressing time-to-event on the clinical-history features. In the AFT framework, the model is parameterized as *log*(*T*) = *μ* + *β^T^X* + *σε* where *β^T^X* is the linear predictor representing the additive shift in log-survival time attributable to the clinical covariates *X*, and *ε* follows either a logistic or standard normal distribution. We extracted this estimated linear predictor 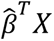 for each individual by applying the coefficient vector 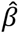, estimated on the training set, to their clinical predictors. For interaction analyses, we used the negative of this linear predictor (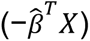) as the continuous effect modifier, so that higher values corresponded to higher predicted disease risk.

### Phenome-wide interaction study in FinnGen and UK Biobank

Among 138 target diseases with available PRS models, we performed two complementary phenome-wide interaction analyses. First, interactions between each disease-specific target PRS and clinical-history phenotypes harmonized between FinnGen and UK Biobank (**Sup. Tab. S1**) were evaluated independently in both cohorts. Second, an extended UK Biobank-only analysis was performed using additional UK Biobank-specific phenotypes, including participant-level and questionnaire-derived traits that could not be harmonized with FinnGen.

For the harmonized analysis, candidate predictors consisted of clinical-history phenotypes recorded before baseline. Phenotypes with an age at onset later than the participant’s baseline age were coded as absent, ensuring that only diagnoses already present at baseline contributed to the analysis.

Phenome-wide interaction analyses were conducted separately for each target disease. Participants who developed the target disease before or at baseline were excluded. Follow-up time was defined as the interval between baseline age and age at target-disease onset or censoring. For each target disease and candidate phenotype, a Cox proportional hazards model was fitted that included baseline age, sex, the disease-specific target PRS, the candidate phenotype, and an interaction term between the PRS and the phenotype. PRSs were adjusted for population structure and standardized to a mean of 0 and a standard deviation of 1 before analysis. The coefficient and the p-value of the interaction term were used to quantify whether the association between the target PRS and disease onset differed according to the candidate phenotype. We restricted analyses to clinical-history phenotypes present in at least 500 individuals, including at least 10 who subsequently developed the target disease.

For the UK Biobank-only analysis, candidate phenotypes were obtained from UK Biobank participant-level and response datasets and processed separately by source dataset. Identifier, demographic, date, age-at-event, target-disease, and target-disease follow-up variables were excluded from the candidate set. Variables were converted to numeric format where applicable, and phenotypes with fewer than 50 non-missing observations or no variation were excluded. Binary variables coded as 0 and 1 were retained on their original scale, whereas non-binary continuous variables were standardized to a mean of 0 and a standard deviation of 1. Each eligible UK Biobank-specific phenotype was then tested for interaction with the disease-specific target PRS using the same Cox model specification as in the harmonized analysis. For continuous phenotypes, models were evaluated only when at least five target-disease cases and five controls were available among participants with complete data.

For sex-specific target diseases, analyses were restricted to participants of the relevant sex, and sex was omitted from the model. Analyses of sex-specific candidate phenotypes were similarly restricted to the corresponding sex. All Cox proportional hazards models were fitted in R using the survival package (v3.4-0).

### Interaction between target disease PRS and predicted clinical risk in FinnGen

For each of the 138 diseases (**Methods: Disease selection**), we assessed whether the hazard ratio associated with the corresponding PRS varied according to an individual’s predicted clinical risk. Clinical risk scores were derived from the corresponding internally and externally validated AFT models, which included age, sex, and clinical-history phenotypes, and were calculated as the sum of predictor values multiplied by their fitted coefficients (**Methods: Calculation of clinical risk on the linear scale for interaction studies**). Clinical risk scores were standardized to a mean of 0 and a standard deviation of 1. The disease-specific PRS, adjusted for population structure, was included as the genetic predictor.

Analyses were restricted to participants with positive follow-up time, and individuals with the target disease before baseline were excluded. For each disease, we fitted a Cox proportional hazards model that included the standardized clinical risk score, the corresponding PRS, baseline age, sex where appropriate for non-sex-specific diseases, and an interaction term between clinical risk and the PRS. In an additional sensitivity analysis, we included PRS-by-age and PRS-by-sex interaction terms in the models. Models were fitted using the survival R package (version 3.4-0), and interaction significance was assessed using Wald tests for the interaction coefficient in Cox proportional hazards models.

Further, we evaluated whether the association between genetic risk and all-cause mortality varied across levels of baseline clinical risk. Clinical risk was represented by the standardized linear predictor from a UK Biobank-trained all-cause mortality model that included age, sex, and clinical-history phenotypes (**Methods. Calculation of clinical risk on linear scale for interaction studies**). Genetic risk was represented by the standardized linear predictor from the corresponding genetic model after removing the contributions of baseline age and sex. Follow-up time was calculated as age at death or censoring minus baseline age. We fitted a Cox proportional hazards model including the main effects of the clinical and genetic linear predictors, their interaction, baseline age, and sex using the survival R package (v3.4-0). The interaction term’s significance was assessed using a Wald test and reflected how the effect of genetic risk on mortality differed according to clinical risk.

### Comparison of the incremental predictive value of genetic information across clinical-risk strata

For each disease, we evaluated whether the improvement in predictive performance obtained by adding genetic information to the clinical model differed according to an individual’s predicted clinical risk (**Methods. Calculation of clinical risk on linear scale for interaction studies**). We grouped individuals into two groups: individuals with scores below and above the median predicted clinical risk for each disease, referred to as the lower-clinical-risk group and the higher-clinical-risk group, respectively. In each group, we measured the performance of clinical and clinical-genetic models using C-index during the follow-up from baseline to disease onset or censoring. Then, the utility of adding genetic information to a clinical model was estimated as the difference in C-index between clinical-genetic and clinical models.

Uncertainty was estimated using 100 bootstrap replicates. Within each clinical-risk stratum, participants with and without an observed event were resampled separately with replacement. In each bootstrap replicate, the C-indices of the clinical and clinical-genetic models, the incremental C-index within each stratum, and the difference in incremental C-index between strata were recalculated.

Wald *P* values for differences in incremental C-index between the lower- and higher-clinical-risk groups were calculated from the estimated differences and their corresponding standard errors and adjusted for multiple testing using the FDR correction. The analysis was restricted to 33 diseases that showed both a significant interaction (**Methods**. **Interaction between target disease PRS and predicted clinical risk in FinnGen**) and a significant genetic increment in tdAUC (**Methods. Comparison of model performance**).

The analysis was repeated independently for each disease endpoint and model specification. Concordance indices were calculated in R using the survival (v3.4-0) package.

### PrognosIQ web application for individualized risk prediction

PrognosIQ was implemented as an interactive R Shiny (v1.10.0) [51] application providing individualized disease-risk estimates from the pretrained disease-specific AFT models described above (**Methods, Genetic and Clinical Genetic Predictive Models; Sup. materials. Additional assessment of model predictive performance and clinical relevance**). We further assessed the calibration and positive predictive value (PPV) of the PrognosIQ models (**Sup. materials. Quality metrics of PrognosIQ models**). Users provide age, sex, smoking status, required clinical measurements, and clinical history. Quantitative predictors are standardized using mean and standard deviation derived from the training data.

For genetic risk estimation, users upload a single-sample VCF or gVCF file in GRCh37 or GRCh38. The application evaluates coverage of score variants and calculates PRS and genetic principal components using PLINK 2.0. PRS are adjusted for population structure by subtracting the component predicted from the genetic principal components and are subsequently standardized relative to the reference population (**Sup. materials. PrognosIQ: polygenic score calculation, harmonization and quality control**). The application then applies the corresponding clinical-genetic model. Cumulative disease risk is estimated annually over 1-10 years and displayed together with the observed population risk for the corresponding disease, sex and baseline-age stratum. Uploaded genotype and intermediate files are deleted after processing.

### External validation of PrognosIQ

PrognosIQ was externally evaluated in an independent prospective cohort previously described by Usoltsev et al. [34]. Participants were recruited in 2012-2013 in 3 Russian metro areas: St. Petersburg, Samara, and Orenburg. Baseline assessment included questionnaires on medical history and lifestyle, anthropometric measurements, and blood biomarkers. Follow-up information on incident diseases and vital status was subsequently obtained through direct participant contact and local healthcare records, with follow-up available through 2022.

PRSs were calculated and adjusted for population structure using the same PCA-based framework as in UK Biobank, with adjustment parameters estimated within Biobank Russia. The adjusted PRSs were then standardized using UK Biobank-derived means and standard deviations to ensure compatibility with the PrognosIQ prediction models (**Sup. materials. Calculation of PRS in Biobank Russia**).

For evaluation of type 2 diabetes prediction, we included 3,644 participants who were free of type 2 diabetes at baseline and had follow-up data available. PrognosIQ predictions were generated using baseline data available in the cohort. During follow-up, 110 participants developed type 2 diabetes. PrognosIQ performance was compared with FINDRISC, which was calculated from baseline age, body mass index, waist circumference, physical activity, fruit and vegetable intake, antihypertensive medication use, history of elevated blood glucose, and family history of diabetes.

ASCVD prediction was evaluated separately in 2,647 participants who were free of cardiovascular disease and were not receiving statin therapy at baseline. During 10 years of follow-up, 86 participants developed ASCVD. PrognosIQ predictions were compared with those obtained using the PREVENT-ASCVD equation. All variables required to calculate FINDRISC and PREVENT-ASCVD were available in the respective validation samples, whereas PrognosIQ predictions were generated using the subset of baseline clinical information available in Biobank Russia (**Sup. materials. PrognosIQ validation in Biobank Russia**).

Within each analysis, PrognosIQ and the corresponding established clinical risk score were evaluated in the same participants. Discrimination was assessed using the area under the receiver operating characteristic curve (ROC AUC), calculated in R using the pROC package (v1.18.5) [52].

### Phenotype analysis of discordant clinical and genetic risk groups

For each disease, the corresponding population-structure-adjusted PRS and clinical-model risk (**Methods. Calculation of clinical risk on linear scale for interaction studies**) were standardized. Participants with prevalent disease at baseline were excluded. We then selected two discordant risk groups: individuals in the top decile of clinical risk and bottom decile of PRS, and individuals in the bottom decile of clinical risk and top decile of PRS. The group indicator was coded as 1 for high clinical risk and low PRS and as 0 for low clinical risk and high PRS.

Each eligible UK Biobank phenotype was tested separately using logistic regression adjusted for baseline age and sex. Continuous phenotypes were standardized, whereas binary phenotypes were analyzed on their original 0/1 scale. Analyses required at least 50 participants, including at least five individuals in each discordant risk group. Associations were reported as odds ratios with 95% confidence intervals and Wald test p-values.

### Visualization

All figures were generated in R using ggplot2 v3.5.2 [53]. Data were imported and prepared for visualization using data.table v1.14.8 [54] and the tidyverse v2.0.0, including dplyr v1.1.4, tidyr v1.3.2, and stringr v1.5.0 [55]. Text labels were positioned using ggrepel v0.9.6 [56], and multipanel figures were assembled using patchwork v1.3.0 and cowplot v1.1.3 [57, 58]. Axis breaks were implemented using ggbreak v0.1.7 [59], scale formatting was performed using scales v1.4.0 [60], and low-level graphical layout was controlled using the R grid v4.3.0 graphics system [61].

## Ethics Statement

FinnGen. Individuals in FinnGen provided informed consent for biobank research, based on the Finnish Biobank Act. Alternatively, separate research cohorts, collected prior to the Finnish Biobank Act came into effect (in September 2013) and the start of FinnGen (August 2017), were collected based on study-specific consents and later transferred to the Finnish biobanks after approval by Fimea (Finnish Medicines Agency) and the National Supervisory Authority for Welfare and Health. Recruitment protocols followed the biobank protocols approved by Fimea. The Coordinating Ethics Committee of the Hospital District of Helsinki and Uusimaa (HUS) statement number for the FinnGen study is Nr HUS/990/2017. Further details on permit and biobank decision numbers are available in the FinnGen Ethics Statement in the Supplementary Information.

UK biobank. This research has been conducted using the UK Biobank Resource under application number 88907. UK Biobank has approval from the North West Multi-centre Research Ethics Committee (MREC) as a Research Tissue Bank (REC reference: 16/NW/0274), and all participants provided written informed consent.

Biobank Russia. All participants signed an informed consent. The study protocol was approved by local ethics committees (Almazov National Medical Research Center, St. Petersburg).

## Data availability

The individual-level FinnGen data used in this study were obtained under FinnGen Proposal Number F_2020_073 (https://www.finngen.fi/en). The FinnGen data are not publicly available because they contain sensitive genetic and health information protected by participant consent, ethical approval, and Finnish and European data protection regulations, including the General Data Protection Regulation (GDPR). Thus, the raw FinnGen data are protected and are not publicly available due to data privacy laws. Details on the data-access restrictions and policies are listed within the FinnGen flagship publication - Kurki et al., Nature, 2023.

This research has been conducted using the UK Biobank Resource under Application Number 88907 (https://www.ukbiobank.ac.uk/). The individual-level UK Biobank data used in this study are available under restricted access because they contain sensitive participant-level genetic and phenotypic information and are governed by UK Biobank access policies. Access can be obtained by bona fide researchers through an application to the UK Biobank. The raw UK Biobank data are protected and are not publicly available due to participant privacy and data governance restrictions.

All PRS models used in this study are publicly available through the PGS Catalog [https://www.pgscatalog.org/] (PGS Catalog) using their corresponding score identifiers.

For Biobank Russia, the open sharing of individual-level clinical and genetic data, including deposition in publicly accessible databases, is not permitted under the laws of the Russian Federation. Therefore, the authors are unable to provide public access to the raw data.

Models developed in this study are available through PrognosIQ, https://prognosiq.nchigm.org, together with tools for calculating individual polygenic risk scores from whole-genome VCF/gVCF files for use as model inputs. Use of PrognosIQ and the predictive models for commercial or for-profit purposes is subject to restrictions associated with provisional patent US PTO 64/138,343. For commercial use or licensing of PrognosIQ, please contact the corresponding author.

## Acknowledgements

We thank the participants and investigators of the FinnGen, the UK Biobank and Biobank Russia.

This research has been conducted using the UK Biobank Resource under Application Number 88907.

A full list of FinnGen Consortium members is shown in the **Sup. Tab. S27**. Following biobanks are acknowledged for delivering biobank samples to FinnGen: Auria Biobank (www.auria.fi/biopankki), THL Biobank (www.thl.fi/biobank), Helsinki Biobank (www.helsinginbiopankki.fi), Biobank Borealis of Northern Finland (https://www.ppshp.fi/Tutkimusja-opetus/Biopankki/Pages/Biobank-Borealis-briefly-in-E), Finnish Clinical Biobank Tampere (www.tays.fi/en-US/Research_and_development/Finnish_Clinical_Biobank_Tampere), Biobank of Eastern Finland (www.itasuomenbiopankki.fi/en), Central Finland Biobank (www.ksshp.fi/fi-FI/Potilaalle/Biopankki), Finnish Red Cross Blood Service Biobank (www.veripalvelu.fi/verenluovutus/biopankkitoiminta), Terveystalo Biobank (www.terveystalo.com/fi/Yritystietoa/Terveystalo-Biopankki/Biopankki/), and Arctic Biobank (https://www.oulu.fi/en/university/faculties-and-units/faculty-medicine/northern-finland-birth-cohorts-andarctic-biobank). All Finnish Biobanks are members of BBMRI.fi infrastructure (www.bbmri.fi). Finnish Biobank Cooperative - FINBB is the coordinator of BBMRI-ERIC operations in Finland. The Finnish biobank data can be accessed through the Fingenious® services (https://site.fingenious.fi/en/) managed by FINBB.

D.U. and M.A. would like to thank the High-Performance Computing core at Abigail Wexner Research Institute for their support of the computational infrastructure for the project.

ChatGPT (GPT-5 and GPT-6) was used during the preparation of this manuscript to improve readability and refine the English language. The authors reviewed and edited all AI-assisted content and take full responsibility for the final text.

## Author contributions

Study design and conceptualization - D.U., I.M., and M.A. Data Analysis - D.U., I.M., and M.A. Statistical model design and manuscript writing D.U., I.M., N.K. and M.A. PrognosIQ application design - D.U., A.U., and M.A. Recruiting patients and collecting biospecimens in Biobank Russia - O.R., An.K., and Al.K. Funding acquisition - M.A., M.J.D., A.P. and S.R. Project supervision - M.A. Manuscript editing and approval - all authors.

## Funding

D.U., I.M., N.K., and M.A. disclose support for this work from the Aging Biology Foundation. A.U. was supported by Nationwide Pediatric Innovations Fund. The FinnGen project is funded by two grants from Business Finland (HUS 4685/31/2016 and UH 4386/31/2016) and by the following industry partners: AbbVie Inc., AstraZeneca UK Ltd, Biogen MA Inc., Bristol Myers Squibb (including Celgene Corporation and Celgene International II Sàrl), Genentech Inc., Merck Sharp & Dohme LLC, Pfizer Inc., GlaxoSmithKline Intellectual Property Development Ltd., Sanofi US Services Inc., Maze Therapeutics Inc., Janssen Biotech Inc., Novartis Pharma AG, and Boehringer Ingelheim International GmbH. UK Biobank is funded by the Medical Research Council, Wellcome, the Department of Health, the Scottish Government, the Welsh Assembly Government, the British Heart Foundation, Cancer Research UK, Diabetes UK, the National Institute for Health and Care Research (NIHR), and the Northwest Regional Development Agency.

## Conflict of interest

A provisional patent has been filed with respect to the findings described in this manuscript (USPTO serial no. 64/138,343). Mark J. Daly is a founder of Maze Therapeutics. The remaining authors declare no competing interests.

## Notes

**Conflict of interest statement is at the end of the manuscript.**

### Author Declarations

FinnGen. Individuals in FinnGen provided informed consent for biobank research, based on the Finnish Biobank Act. Alternatively, separate research cohorts, collected prior to the Finnish Biobank Act came into effect (in September 2013) and the start of FinnGen (August 2017), were collected based on study-specific consents and later transferred to the Finnish biobanks after approval by Fimea (Finnish Medicines Agency) and the National Supervisory Authority for Welfare and Health. Recruitment protocols followed the biobank protocols approved by Fimea. The Coordinating Ethics Committee of the Hospital District of Helsinki and Uusimaa (HUS) statement number for the FinnGen study is Nr HUS/990/2017. Further details on permit and biobank decision numbers are available in the FinnGen Ethics Statement in the Supplementary Information. UK biobank. This research has been conducted using the UK Biobank Resource under application number 88907. UK Biobank has approval from the North West Multi-centre Research Ethics Committee (MREC) as a Research Tissue Bank (REC reference: 16/NW/0274), and all participants provided written informed consent. Biobank Russia. All participants signed an informed consent. The study protocol was approved by local ethics committees (Almazov National Medical Research Center, St. Petersburg)

## References

1. Polygenic Risk Score Task Force of the International Common Disease Alliance. Responsible use of polygenic risk scores in the clinic: potential benefits, risks and gaps. Nat Med 27, 1876–1884 (2021). 10.1038/s41591-021-01549-6

2. Kullo, I.J., Lewis, C.M., Inouye, M. et al. Polygenic scores in biomedical research. Nat Rev Genet 23, 524–532 (2022). 10.1038/s41576-022-00470-z

3. Kolosov, N., Reeve, M.P., Briotta Parolo, P.D. et al. PGS Browser: a public platform for personalized polygenic score analysis and interpretation. Nat Commun (2026). 10.1038/s41467-026-74461-7

4. Urbut, S.M., Ding, Y., Nakao, T. et al. A Bayesian framework for longitudinal EHR and genetic discovery. Nature (2026). 10.1038/s41586-026-10780-5

5. Chatterjee N, Shi J, García-Closas M. Developing and evaluating polygenic risk prediction models for stratified disease prevention. Nat Rev Genet. 2016 Jul;17(7):392–406. doi: 10.1038/nrg.2016.27.

6. Lewis ACF, Green RC, Vassy JL. Polygenic risk scores in the clinic: Translating risk into action. HGG Adv. 2021 Jul 28;2(4):100047. doi: 10.1016/j.xhgg.2021.100047.

7. Abu-El-Haija A, Reddi HV, Wand H, Rose NC, Mori M, Qian E, Murray MF; ACMG Professional Practice and Guidelines Committee. The clinical application of polygenic risk scores: A points to consider statement of the American College of Medical Genetics and Genomics (ACMG). Genet Med. 2023 May;25(5):100803. doi: 10.1016/j.gim.2023.100803.

8. Sinnott-Armstrong, N., Tanigawa, Y., Amar, D. et al. Genetics of 35 blood and urine biomarkers in the UK Biobank. Nat Genet 53, 185–194 (2021). 10.1038/s41588-020-00757-z

9. Mosley JD, Gupta DK, Tan J, Yao J, Wells QS, Shaffer CM, Kundu S, Robinson-Cohen C, Psaty BM, Rich SS, Post WS, Guo X, Rotter JI, Roden DM, Gerszten RE, Wang TJ. Predictive Accuracy of a Polygenic Risk Score Compared With a Clinical Risk Score for Incident Coronary Heart Disease. JAMA. 2020 Feb 18;323(7):627–635. doi: 10.1001/jama.2019.21782.

10. Elliott J, Bodinier B, Bond TA, et al. Predictive Accuracy of a Polygenic Risk Score– Enhanced Prediction Model vs a Clinical Risk Score for Coronary Artery Disease. JAMA. 2020;323(7):636–645. doi:10.1001/jama.2019.22241

11. Mars N, Koskela JT, Ripatti P, Kiiskinen TTJ, Havulinna AS, Lindbohm JV, Ahola-Olli A, Kurki M, Karjalainen J, Palta P; FinnGen; Neale BM, Daly M, Salomaa V, Palotie A, Widén E, Ripatti S. Polygenic and clinical risk scores and their impact on age at onset and prediction of cardiometabolic diseases and common cancers. Nat Med. 2020 Apr;26(4):549–557. doi: 10.1038/s41591-020-0800-0.

12. Lee A, Mavaddat N, Wilcox AN, Cunningham AP, Carver T, Hartley S, Babb de Villiers C, Izquierdo A, Simard J, Schmidt MK, Walter FM, Chatterjee N, Garcia-Closas M, Tischkowitz M, Pharoah P, Easton DF, Antoniou AC. BOADICEA: a comprehensive breast cancer risk prediction model incorporating genetic and nongenetic risk factors. Genet Med. 2019 Aug;21(8):1708–1718. doi: 10.1038/s41436-018-0406-9. Epub 2019 Jan 15. Erratum in: Genet Med. 2019 Jun;21(6):1462. doi: 10.1038/s41436-019-0459-4.

13. Martin AR, Kanai M, Kamatani Y, Okada Y, Neale BM, Daly MJ. Clinical use of current polygenic risk scores may exacerbate health disparities. Nat Genet. 2019 Apr;51(4):584–591. doi: 10.1038/s41588-019-0379-x. Epub 2019 Mar 29. Erratum in: Nat Genet. 2021 May;53(5):763. doi: 10.1038/s41588-021-00797-z. PMID: 30926966; PMCID: PMC6563838.

14. Duncan, L., Shen, H., Gelaye, B. et al. Analysis of polygenic risk score usage and performance in diverse human populations. Nat Commun 10, 3328 (2019). 10.1038/s41467-019-11112-0

15. Kachuri, L., Chatterjee, N., Hirbo, J. et al. Principles and methods for transferring polygenic risk scores across global populations. Nat Rev Genet 25, 8–25 (2024). 10.1038/s41576-023-00637-2

16. Wang, Y., Guo, J., Ni, G. et al. Theoretical and empirical quantification of the accuracy of polygenic scores in ancestry divergent populations. Nat Commun 11, 3865 (2020). 10.1038/s41467-020-17719-y

17. Mostafavi H, Harpak A, Agarwal I, Conley D, Pritchard JK, Przeworski M. Variable prediction accuracy of polygenic scores within an ancestry group. Elife. 2020 Jan 30;9:e48376. doi: 10.7554/eLife.48376.

18. Ye Y, Chen X, Han J, Jiang W, Natarajan P, Zhao H. Interactions Between Enhanced Polygenic Risk Scores and Lifestyle for Cardiovascular Disease, Diabetes, and Lipid Levels. Circ Genom Precis Med. 2021 Feb;14(1):e003128. doi: 10.1161/CIRCGEN.120.003128.

19. Tanigawa Y, Qian J, Venkataraman G, Justesen JM, Li R, Tibshirani R, Hastie T, Rivas MA. Significant sparse polygenic risk scores across 813 traits in UK Biobank. PLoS Genet. 2022 Mar 24;18(3):e1010105. doi: 10.1371/journal.pgen.1010105.

20. Nagpal, S., Gibson, G. Pervasive interactions between exposures and polygenic risk can inform more effective clinical and behavioral interventions. Nat Genet (2026). 10.1038/s41588-026-02674-z

21. Said MA, Verweij N, van der Harst P. Associations of Combined Genetic and Lifestyle Risks With Incident Cardiovascular Disease and Diabetes in the UK Biobank Study. JAMA Cardiol. 2018 Aug 1;3(8):693–702. doi: 10.1001/jamacardio.2018.1717.

22. Dashti HS, Miranda N, Cade BE, Huang T, Redline S, Karlson EW, Saxena R. Interaction of obesity polygenic score with lifestyle risk factors in an electronic health record biobank. BMC Med. 2022 Jan 12;20(1):5. doi: 10.1186/s12916-021-02198-9.

23. Bycroft C, Freeman C, Petkova D, Band G, Elliott LT, Sharp K, Motyer A, Vukcevic D, Delaneau O, O’Connell J, Cortes A, Welsh S, Young A, Effingham M, McVean G, Leslie S, Allen N, Donnelly P, Marchini J. The UK Biobank resource with deep phenotyping and genomic data. Nature. 2018 Oct;562(7726):203–209. doi: 10.1038/s41586-018-0579-z. Epub 2018 Oct 10. PMID: 30305743; PMCID: PMC6786975.

24. Kurki, M.I., Karjalainen, J., Palta, P. et al. FinnGen provides genetic insights from a well-phenotyped isolated population. Nature 613, 508–518 (2023). 10.1038/s41586-022-05473-8.

25. Wei LJ. The accelerated failure time model: a useful alternative to the Cox regression model in survival analysis. Statistics in Medicine. 1992;11:1871–1879. doi:10.1002/sim.4780111409.

26. Zou H, Hastie T. Regularization and variable selection via the elastic net. Journal of the Royal Statistical Society: Series B. 2005;67:301–320. doi:10.1111/j.1467-9868.2005.00503.x

27. Heagerty PJ, Lumley T, Pepe MS. Time-dependent ROC curves for censored survival data and a diagnostic marker. Biometrics. 2000;56:337–344. doi:10.1111/j.0006-341X.2000.00337.x

28. Detrois, K.E., Hartonen, T., Teder-Laving, M. et al. Cross-biobank generalizability and accuracy of electronic health record-based predictors compared to polygenic scores. Nat Genet 57, 2136–2145 (2025). 10.1038/s41588-025-02298-9

29. Khan SS, Matsushita K, Sang Y, et al. Development and Validation of the American Heart Association’s PREVENT Equations. Circulation. 2024;149(6):430–449. doi:10.1161/CIRCULATIONAHA.123.067626.

30. Lindström J, Tuomilehto J. The diabetes risk score: a practical tool to predict type 2 diabetes risk. Diabetes Care. 2003 Mar;26(3):725–31. doi: 10.2337/diacare.26.3.725. PMID: 12610029.

31. Truong B, Hull LE, Ruan Y, Huang QQ, Hornsby W, Martin H, van Heel DA, Wang Y, Martin AR, Lee SH, Natarajan P. Integrative polygenic risk score improves the prediction accuracy of complex traits and diseases. Cell Genom. 2024 Apr 10;4(4):100523. doi: 10.1016/j.xgen.2024.100523.

32. Blumenthal RS, Morris PB, Gaudino M, et al. 2026 ACC/AHA/AACVPR/ABC/ACPM/ADA/AGS/APhA/ASPC/NLA/PCNA Guideline on the Management of Dyslipidemia: A Report of the American College of Cardiology/American Heart Association Joint Committee on Clinical Practice Guidelines. J Am Coll Cardiol. 2026;87(19):2624–2757. doi: 10.1016/j.jacc.2025.11.016.

33. Miao, J., Song, G., Wu, Y. et al. PIGEON: a statistical framework for estimating gene–environment interaction for polygenic traits. Nat Hum Behav 9, 1654–1668 (2025). 10.1038/s41562-025-02202-9

34. Usoltsev, D., Kolosov, N., Rotar, O. et al. Complex trait susceptibilities and population diversity in a sample of 4,145 Russians. Nat Commun 15, 6212 (2024). 10.1038/s41467-024-50304-1

35. Manuel A.R. Ferreira, Riddhima Mathur, Judith M. Vonk, Agnieszka Szwajda, Ben Brumpton, Raquel Granell, Bronwyn K. Brew, Vilhelmina Ullemar, Yi Lu, Yunxuan Jiang, Patrik K.E. Magnusson, Robert Karlsson, David A. Hinds, Lavinia Paternoster, Gerard H. Koppelman, Catarina Almqvist,Genetic Architectures of Childhood- and Adult-Onset Asthma Are Partly Distinct,The American Journal of Human Genetics,Volume 104, Issue 4, 2019,Pages 665–684, 10.1016/j.ajhg.2019.02.022.

36. Khera, A.V., Chaffin, M., Aragam, K.G. et al. Genome-wide polygenic scores for common diseases identify individuals with risk equivalent to monogenic mutations. Nat Genet 50, 1219–1224 (2018). 10.1038/s41588-018-0183-z

37. Inouye M, Abraham G, Nelson CP, Wood AM, Sweeting MJ, Dudbridge F, Lai FY, Kaptoge S, Brozynska M, Wang T, Ye S, Webb TR, Rutter MK, Tzoulaki I, Patel RS, Loos RJF, Keavney B, Hemingway H, Thompson J, Watkins H, Deloukas P, Di Angelantonio E, Butterworth AS, Danesh J, Samani NJ; UK Biobank CardioMetabolic Consortium CHD Working Group. Genomic Risk Prediction of Coronary Artery Disease in 480,000 Adults: Implications for Primary Prevention. J Am Coll Cardiol. 2018 Oct 16;72(16):1883–1893. doi: 10.1016/j.jacc.2018.07.079.

38. Kochanek KD, Murphy SL, Xu JQ, Arias E. Mortality in the United States, 2022. NCHS Data Brief, no. 492. National Center for Health Statistics; 2024. 10.15620/cdc:135850

39. Therneau TM, Grambsch PM. Modeling Survival Data: Extending the Cox Model. New York: Springer; 2000.

40. Therneau TM. A Package for Survival Analysis in R. R package version 3.4-0. 2022. Available from: https://CRAN.R-project.org/package=survival.

41. McFadden D. Conditional logit analysis of qualitative choice behavior. In: Zarembka P, editor. Frontiers in Econometrics. New York: Academic Press; 1974. p. 105–142.

42. Davidson-Pilon C. lifelines: survival analysis in Python. Journal of Open Source Software. 2019;4(40):1317. doi:10.21105/joss.01317.

43. Virtanen P, Gommers R, Oliphant TE, et al. SciPy 1.0: fundamental algorithms for scientific computing in Python. Nature Methods. 2020;17:261–272. doi:10.1038/s41592-019-0686-2.

44. Blanche P, Dartigues JF, Jacqmin-Gadda H. Estimating and comparing time-dependent areas under receiver operating characteristic curves for censored event times with competing risks. Statistics in Medicine. 2013;32(30):5381–5397. doi:10.1002/sim.5958.

45. Jaeger, B. C. PooledCohort: Predicted Risk for CVD Using Pooled Cohort Equations, PREVENT Equations, and Other Contemporary CVD Risk Calculators. R package version 0.0.2 (2024). https://CRAN.R-project.org/package=PooledCohort

46. Inker LA, Eneanya ND, Coresh J, et al. New creatinine- and cystatin C–based equations to estimate GFR without race. N Engl J Med. 2021;385:1737–1749. doi:10.1056/NEJMoa2102953.

47. Lambert SA, Gil L, Jupp S, et al. The Polygenic Score Catalog as an open database for reproducibility and systematic evaluation. Nature Genetics. 2021;53:420–425. doi:10.1038/s41588-021-00783-5.

48. Lambert SA, Wingfield B, Gibson JT, et al. Enhancing the Polygenic Score Catalog with tools for score calculation and ancestry normalization. Nature Genetics. 2024;56:1989–1994. doi:10.1038/s41588-024-01937-x

49. Chang CC, Chow CC, Tellier LCAM, Vattikuti S, Purcell SM, Lee JJ. Second-generation PLINK: rising to the challenge of larger and richer datasets. GigaScience. 2015;4:7. doi:10.1186/s13742-015-0047-8.

50. Weissbrod, O., Kanai, M., Shi, H. et al. Leveraging fine-mapping and multipopulation training data to improve cross-population polygenic risk scores. Nat Genet 54, 450–458 (2022). 10.1038/s41588-022-01036-9

51. Chang W, Cheng J, Allaire JJ, Sievert C, Schloerke B, Xie Y, Allen J, McPherson J, Dipert A, Borges B. shiny: Web Application Framework for R. R package version 1.10.0. 2024. Available from: https://CRAN.R-project.org/package=shiny

52. Robin X, Turck N, Hainard A, Tiberti N, Lisacek F, Sanchez JC, Müller M. pROC: an open-source package for R and S+ to analyze and compare ROC curves. BMC Bioinformatics. 2011;12:77. doi:10.1186/1471-2105-12-77

53. Wickham H. ggplot2: Elegant Graphics for Data Analysis. New York: Springer-Verlag; 2016.

54. Barrett T, Dowle M, Srinivasan A, et al. data.table: Extension of data.frame. R package.

55. Wickham H, Averick M, Bryan J, et al. Welcome to the tidyverse. Journal of Open Source Software. 2019;4(43):1686. doi:10.21105/joss.01686.

56. Slowikowski K. ggrepel: Automatically Position Non-Overlapping Text Labels with ggplot2. R package.

57. Pedersen TL. patchwork: The Composer of Plots. R package.

58. Wilke CO. cowplot: Streamlined Plot Theme and Plot Annotations for ggplot2. R package.

59. Xu S, Chen M, Feng T, Zhan L, Zhou L, Yu G. Use ggbreak to effectively utilize plotting space to deal with large datasets and outliers. Frontiers in Genetics. 2021;12:774846. doi:10.3389/fgene.2021.774846.

60. Wickham H, Pedersen TL, Seidel D. scales: Scale Functions for Visualization. R package.

61. R Core Team (2023). R: A language and environment for statistical computing. R Foundation for Statistical Computing, Vienna, Austria. https://www.R-project.org/

