## Supplementary Materials for "Clinical history shapes the predictive value of polygenic risk"

### Supplementary Notes

**Table of contents:**

**Ethics Statement** ... 3
**Harmonization of UK Biobank clinical-history phenotypes with FinnGen endpoint definitions** ... 4
**Harmonization of laboratory measurements between FinnGen and UK Biobank** ... 5
**Calculation of PRS in UK Biobank** ... 6
**Additional assessment of model predictive performance and clinical relevance** ... 7
**PrognosIQ web application** ... 8
  **Quality metrics of PrognosIQ models** ... 8
**PrognosIQ: WGS-based polygenic score calculation, harmonization, and quality control** 9
  **Variant preprocessing and harmonization** ... 10
  **PRS calculation** ... 11
  **Population structure adjustment** ... 11
**Calculation of PRS in Biobank Russia** ... 12
**PrognosIQ validation in Biobank Russia** ... 12
**Supplementary figures** ... 14
**References** ... 24

### Ethics Statement

Ethics statement and materials & methods: Study subjects in FinnGen provided informed consent for biobank research, based on the Finnish Biobank Act. Alternatively, separate research cohorts, collected prior the Finnish Biobank Act came into effect (in September 2013) and start of FinnGen (August 2017), were collected based on study-specific consents and later transferred to the Finnish biobanks after approval by Fimea (Finnish Medicines Agency), the National Supervisory Authority for Welfare and Health. Recruitment protocols followed the biobank protocols approved by Fimea. The Coordinating Ethics Committee of the Hospital District of Helsinki and Uusimaa (HUS) statement number for the FinnGen study is Nr HUS/990/2017. The FinnGen study is approved by Finnish Institute for Health and Welfare (permit numbers: THL/2031/6.02.00/2017, THL/1101/5.05.00/2017, THL/341/6.02.00/2018, THL/2222/6.02.00/2018, THL/283/6.02.00/2019, THL/1721/5.05.00/2019 and THL/1524/5.05.00/2020), Digital and population data service agency (permit numbers: VRK43431/2017-3, VRK/6909/2018-3, VRK/4415/2019-3), the Social Insurance Institution (permit numbers: KELA 58/522/2017, KELA 131/522/2018, KELA 70/522/2019, KELA 98/522/2019, KELA 134/522/2019, KELA 138/522/2019, KELA 2/522/2020, KELA 16/522/2020), Findata permit numbers THL/2364/14.02/2020, THL/4055/14.06.00/2020, THL/3433/14.06.00/2020, THL/4432/14.06/2020, THL/5189/14.06/2020, THL/5894/14.06.00/2020, THL/6619/14.06.00/2020, THL/209/14.06.00/2021, THL/688/14.06.00/2021, THL/1284/14.06.00/2021, THL/1965/14.06.00/2021, THL/5546/14.02.00/2020, THL/2658/14.06.00/2021, THL/4235/14.06.00/2021, Statistics Finland (permit numbers: TK-53-1041-17 and TK/143/07.03.00/2020 (earlier TK-53-90-20) TK/1735/07.03.00/2021, TK/3112/07.03.00/2021) and Finnish Registry for Kidney Diseases permission/extract from the meeting minutes on 4th July 2019. The Biobank Access Decisions for FinnGen samples and data utilized in FinnGen Data Freeze 11 include: THL Biobank BB2017_55, BB2017_111, BB2018_19, BB_2018_34, BB_2018_67, BB2018_71, BB2019_7, BB2019_8, BB2019_26, BB2020_1, BB2021_65, Finnish Red Cross Blood Service Biobank 7.12.2017, Helsinki Biobank HUS/359/2017, HUS/248/2020, HUS/430/2021 §28, §29, HUS/150/2022 §12, §13, §14, §15, §16, §17, §18, §23, §58 and §59, Auria Biobank AB17-5154 and amendment #1 (August 17 2020) and amendments BB_2021-0140, BB_2021-0156 (August 26 2021, Feb 2 2022), BB_2021-0169, BB_2021-0179, BB_2021-0161, AB20-5926 and amendment #1 (April 23 2020) and it´s modification (Sep 22 2021), BB_2022-0262, BB_2022-0256, Biobank Borealis of Northern Finland_2017_1013, 2021_5010, 2021_5018, 2021_5015, 2021_5015 Amendment, 2021_5023, 2021_5023 Amendment, 2021_5017, 2022_6001, 2022_6006 Amendment, BB22- 0067, 2022_0262, Biobank of Eastern Finland 1186/2018 and amendment 22§/2020, 53§/2021, 13§/2022, 14§/2022, 15§/2022, 27§/2022, 28§/2022, 29§/2022, 33§/2022, 35§/2022, 36§/2022, 37§/2022, 39§/2022,

7§/2023, Finnish Clinical Biobank Tampere MH0004 and amendments (21.02.2020 & 06.10.2020), 8§/2021, 9§/2021, §9/2022, §10/2022, §12/2022, 13§/2022, §20/2022, §21/2022, §22/2022, §23/2022, 28§/2022, 29§/2022, 30§/2022, 31§/2022, 32§/2022, 38§/2022, 40§/2022, 42§/2022, 1§/2023, Central Finland Biobank 1-2017, BB_2021-0161, BB_2021-0169, BB_2021-0179, BB_2021-0170, BB_2022-0256, and Terveystalo Biobank STB 2018001 and amendment 25th Aug 2020, Finnish Hematological Registry and Clinical Biobank decision 18th June 2021, Arctic biobank P0844: ARC_2021_1001.

### Harmonization of UK Biobank clinical-history phenotypes with FinnGen endpoint definitions

To harmonize clinical-history phenotype definitions between UK Biobank and FinnGen, ICD-10 definitions for FinnGen clinical endpoints were obtained using the FinnGen phenotype-matching pipeline (https://github.com/FINNGEN/phenotype-matching). The FINNGEN_ENDPOINTS_DF11_Final_2022-10-05_public.tsv file was used as the reference source for FinnGen endpoint definitions. For each FinnGen endpoint, the resulting mapping table contained the ICD-10 codes identified by the phenotype-matching pipeline together with the corresponding endpoint-specific regular-expression pattern.

The mapping table was then applied to the ICD-10 diagnoses available in UK Biobank. Directly matched and regular-expression-derived codes were combined to define the final set of ICD-10 diagnoses representing each FinnGen endpoint. Endpoints for which no suitable ICD-10 match was identified were excluded from further processing.

ICD-10 diagnoses obtained from UK Biobank hospital inpatient records (fields 41270 and 41280) and UK Biobank-derived first-occurrence fields (fields 130000-132604), which integrate information from hospital admissions, primary care, death records, and self-reported conditions, were combined with diagnoses from the UK Biobank cancer registry (fields 40006 and 40008). This allowed diagnoses captured through the cancer registry and other linked data sources to contribute to the corresponding FinnGen endpoint definitions (**Sup. Tab. S1**).

At the participant level, a FinnGen endpoint was considered present when at least one of its constituent qualifying diagnoses was recorded. When multiple ICD-10 diagnoses or subcodes contributed to the same endpoint, the earliest recorded age among the qualifying diagnoses was used as the endpoint onset age. Consequently, multiple UK Biobank diagnostic records corresponding to the same FinnGen endpoint were collapsed into a single participant-level phenotype with a binary case indicator and an associated age at first qualifying diagnosis.

Following the initial ICD-10-based construction of FinnGen phenotypes, endpoint definitions were further refined according to the additional case and control criteria specified in the FinnGen endpoint definition file (https://www.finngen.fi/en/researchers/clinical-endpoints, DF11). For endpoints with additional CONDITIONS, candidate cases were retained only when the required accompanying phenotype was present with the appropriate temporal relationship to the endpoint onset; corresponding CONTROL_CONDITIONS and CONTROL_EXCLUDE criteria were applied to restrict the eligible control population. Endpoint-specific rules that could not be represented by direct ICD-10 mapping were implemented separately, including age-at-onset criteria, relationships between related diagnoses, and additional clinical criteria. Sex-specific endpoint definitions were restricted to participants of the applicable sex, whereas participants of the other sex were treated as ineligible for that endpoint.

Phenotypes with CONDITIONS or CONTROL_CONDITIONS in the FinnGen endpoint definitions were excluded from subsequent feature-selection procedures used for model development because, in FinnGen, the phenotypes specified in these criteria were not necessarily required to occur before the corresponding disease endpoint and could therefore arise after disease onset.

#### Harmonization of laboratory measurements between FinnGen and UK Biobank

For FinnGen participants, all available laboratory measurements obtained before baseline age were collected, and the mean value of each laboratory measurement was calculated for each participant. FinnGen laboratory measurements were then mapped to the corresponding UK Biobank laboratory phenotypes. To reduce the influence of extreme values, measurements outside six standard deviations from the FinnGen cohort mean were treated as missing. Where necessary, laboratory values were rescaled to ensure consistency with the corresponding UK Biobank measurements. Missing values were imputed using the corresponding UK Biobank mean values, and laboratory measurements were subsequently standardized using the UK Biobank phenotype-specific mean and standard deviation used for model development (**Sup. Tab. S6**).

#### Analysis of time-dependent features

To characterize common temporal patterns of associations between clinical-history phenotypes and mortality, the effect estimates obtained from the time-dependent Cox models were additionally subjected to clustering. Clustering was performed separately within each age group and biobank. For each clinical-history phenotype, the profile consisted of the estimated log hazard ratios across the six time-since-onset intervals (0-1, 1-3, 3-5, 5-7, 7-10, and >10 years) together with their corresponding standard errors.

We applied a Gaussian mixture model with three clusters. In contrast to conventional clustering based only on point estimates, this approach incorporated the uncertainty of each Cox regression coefficient. Specifically, for phenotype (i), time interval (j), and cluster (k), the observed coefficient was modeled as:

$$\hat{\beta}_{ij} \mid z_i = k \sim N(\mu_{kj}, \tau_{kj}^2 + SE_{ij}^2),$$

where $\mu_{kj}$ represents the cluster-specific mean coefficient, $\tau_{kj}^2$ represents between-phenotype variability within the cluster, and $SE_{ij}^2$ represents the sampling variance of the individual effect estimate. Thus, coefficients with larger standard errors had a smaller influence on cluster assignment.

Before clustering, coefficients and their standard errors were scaled according to the variability of the coefficients within each time interval. Initial cluster assignments were obtained using k-means clustering with 20 random starts. Cluster membership probabilities, cluster-specific mean profiles, within-cluster variances, and mixing proportions were subsequently estimated iteratively until convergence using expectation-maximization algorithm. Each clinical-history phenotype was assigned to the cluster with the highest posterior probability. Clusters were subsequently ordered according to the estimated effect in the earliest available time-since-onset interval to facilitate comparison across age groups and biobanks. Only coefficient-standard error pairs with finite values and standard errors >0 and <10 were included, and at least three available time intervals were required for clustering within a given stratum.

### Calculation of PRS in UK Biobank

UK Biobank imputed genotype data (Field 22828) were used for PRS calculation and population-structure adjustment. Genotype imputation was performed by UK Biobank using the Haplotype Reference Consortium (HRC) and the combined UK10K/1000 Genomes Phase 3 reference panels, as previously described by Bycroft et al. [1]. For both PRS calculation and principal-component projection, analyses were restricted to imputed variants with an INFO score ≥0.8. A total of 373 unique PRS models used as features in the genetic and clinical-genetic prediction models were retrieved from the PGS Catalog using PGS Catalog utilities [2,3]. PRS scoring files were harmonized to the GRCh37 genome build and matched to the filtered UK Biobank imputed variant set. A minimum overlap of 75% between variants included in each PRS and variants available in UK Biobank was required for matching, and all 373 PRS models met this criterion. Individual-level PRSs were calculated using PLINK2 --score [4] by applying the corresponding PGS Catalog variant weights to imputed genotype dosages. Weighted score contributions were calculated separately for each autosome (chromosomes 1-22) and subsequently summed across chromosomes to obtain a PRS for each participant and PRS model. Each PRS was normalized by twice the number of unique matched variants contributing to the score.

To account for population structure, the filtered UK Biobank imputed genotype data were projected onto principal-component axes derived from the 1000 Genomes Project reference panel using HapMap3 variants [5]. Projection was performed in PLINK2 using precomputed 1000 Genomes allele-specific PCA loadings, with genotype dosages variance-standardized according to the reference allele frequencies. Each normalized PRS was then regressed on the first six projected genetic principal components (PC1-PC6), and the fitted PC-associated component was subtracted from the normalized PRS, following previously described ancestry-adjustment approaches [6]. The resulting residualized PRS was subsequently z-standardized separately within the UK Biobank and used in downstream analyses.

#### Additional assessment of model predictive performance and clinical relevance

We characterized the overall predictive performance of the null, clinical, genetic, and clinical-genetic models across the evaluated outcomes (**Sup. Tab. S12**). Relative to the null model containing age and sex, either the clinical or clinical-genetic model significantly improved prediction for 132 outcomes, including all-cause mortality. Significant positive increments were observed for 119 clinical models and for 131 clinical-genetic models. To summarize absolute predictive performance across disease classes, these 132 outcomes were assigned to 16 functional categories, and up to three diseases with the highest mean 1-5-year tdROC AUC were selected for visualization within each category; all prespecified common causes of death were retained (**Sup. Fig. S2**).

Absolute predictive performance varied substantially across disease groups (**Sup. Fig. S2, Sup. Tab. S15**). The clinical-genetic model for all-cause mortality achieved mean 1-5-year tdROC AUC (0.822; 95% CI, 0.817-0.825). Among disease categories, the highest mean performance was observed for common causes of death (0.795 ± 0.019), followed by metabolic and endocrine diseases (0.775 ± 0.023), neurological diseases (0.757 ± 0.032), behavioral disorders (0.743 ± 0.038), and cancer (0.727 ± 0.016).

Although integration of clinical and genetic information generally yielded the strongest prediction, genetic-only models achieved higher tdROC AUC than the corresponding clinical-genetic models for 18 diseases, with a mean difference of 0.014 (**Sup. Tab. S16**). These results demonstrate that the relative contribution of clinical and genetic information varies considerably across outcomes, even when both sources of information are individually predictive.

To determine whether the predictive gains from genetic information translated into clinically meaningful changes in risk classification, we compared UK Biobank participants who crossed the 5% 10-year ASCVD risk threshold in either direction after genetics was incorporated into the clinical model (**Sup. Fig. S5**). In addition to the 4,178 participants reclassified upward from <5% to ≥5%, 1,186 were reclassified downward from ≥5% to <5%. Thus, upward reclassification occurred approximately 3.5 times more frequently than downward reclassification. This directional asymmetry indicates that the principal effect of incorporating genetic information was to identify additional individuals whose risk was not captured by the clinical model, rather than to remove individuals already classified as high risk. Together with the substantially elevated observed incidence among upward-reclassified participants, these findings support a potential role for genetic information in identifying clinically relevant risk among individuals whose conventional clinical profiles would otherwise place them below an intervention threshold.

### PrognosIQ web application

PrognosIQ (https://prognosiq.nchigm.org) is a web-based application that provides individualized estimates of absolute disease risk using the disease-specific accelerated failure time (AFT) models developed in UK Biobank (**Methods. Genetic and clinical genetic predictive models**). The application implements clinical-genetic models that integrate medical history, routine biomarkers, and polygenic risk scores (PRS). These models generate risk estimates over 10-years prediction horizons from the individual's clinical and genetic profile.

### Quality metrics of PrognosIQ models

The predictive performance of PrognosIQ models was evaluated using complementary measures of discrimination, calibration, and clinical utility. External discrimination was assessed among participants aged 40-70 years in the independent FinnGen cohort, whereas calibration of absolute risk estimates was evaluated in the held-out UK Biobank validation cohort. For each endpoint, the best-performing model was selected from the genetic, clinical, and clinical-genetic models. Models were included in PrognosIQ if their mean 1-5-year tdROC AUC exceeded 0.70 in the independent FinnGen validation cohort and they showed a statistically significant improvement over the corresponding null model (two-sided Wald test, FDR-adjusted $P < 0.05$). 69 models met both criteria.

In addition to tdROC AUC (**Methods. Estimation of models performance**), model performance was characterized using the Brier score, which evaluates the accuracy of probabilistic predictions by jointly reflecting discrimination and calibration. The 5-year Brier score was calculated using an inverse-probability-of-censoring approach implemented in the pec R package v2025.6.24 [7], with the censoring distribution modeled using a Cox proportional hazards model. Confidence intervals were obtained by nonparametric bootstrap resampling using 100 bootstrap replicates, with 95% confidence intervals defined by the 2.5th and 97.5th percentiles of the valid bootstrap estimates. The same bootstrap samples were used across aligned models to facilitate direct comparisons between models.

Calibration of 5-year absolute risk predictions was assessed in the UK Biobank validation cohort by comparing predicted and observed event probabilities. Overall observed 5-year risk was estimated as 1−S(5), where S(5) was obtained using the Kaplan-Meier estimator, and was compared with the mean predicted 5-year risk. Calibration-in-the-large was summarized using the observed-to-expected (O/E) risk ratio, with a value of 1 indicating agreement between predicted and observed risk. Calibration intercept and calibration slope were additionally estimated at the 5-year horizon using inverse-probability-of-censoring-weighted logistic regression to account for incomplete follow-up. For calibration intercept estimation, the logit-transformed predicted risk was included as an offset, such that an intercept of 0 indicates ideal overall calibration. Calibration slope was estimated by regressing the censoring-adjusted 5-year outcome on the logit-transformed predicted risk; a slope of 1 indicates ideal calibration, values below 1 indicate predictions that are too extreme, and values above 1 indicate predictions that are insufficiently dispersed. Calibration curves were generated by stratifying individuals into ten groups according to predicted 5-year risk and plotting the mean predicted risk within each group against the corresponding Kaplan-Meier estimate of observed 5-year risk, with the identity line representing perfect calibration.

Clinical utility was additionally assessed using the positive predictive value (PPV) at prespecified predicted-risk thresholds of 0.01%, 0.1%, 1%, 3%, 5%, 10%, 20%, and 50%. For each threshold, individuals with a predicted risk equal to or greater than the corresponding cutoff were classified as test-positive. Because follow-up was subject to censoring, PPV at 5 years was estimated as 1−S(5), where S(5) represents the Kaplan-Meier estimate of event-free survival at 5 years among individuals classified as test-positive. Confidence intervals for PPV were derived from the corresponding Kaplan-Meier confidence limits (**Sup. Tab. S25**)

### PrognosIQ: WGS-based polygenic score calculation, harmonization, and quality control

Polygenic risk scores (PRSs) are calculated in PrognosIQ directly from individual-level whole-genome sequencing (WGS) data provided in VCF or gVCF format. The workflow is designed for genome-wide variant data generated by WGS and performs build-specific preprocessing, harmonization, quality control, and PRS calculation. Both GRCh37/hg19 and GRCh38/hg38 WGS inputs are supported. The genome build is determined from assembly annotations in the VCF header or, when these are unavailable, from chromosome contig lengths. Users can also specify the genome build manually; discordance between the selected and automatically detected build terminates the calculation. Separate reference FASTA files, target-site sets, allele-frequency resources, PCA reference files, and PRS score files are used for WGS data aligned to GRCh37 and GRCh38. No liftover between genome builds is performed during the scoring workflow.

### Variant preprocessing and harmonization

WGS VCF files are automatically bgzip-compressed and indexed by the application when required for downstream processing. Multiallelic records are decomposed using BCFtools v1.23 [8], exact duplicate variants are removed, and the genome-wide variant data are restricted to the predefined set of sites represented in the build-specific scoring resources. Variant identifiers are standardized to the CHROM_POS_REF_ALT format before downstream processing.

For WGS data supplied in gVCF format, genotypes are generated using GATK v4.6.2.0 GenotypeGVCFs [9] with the corresponding whole-genome reference assembly and predefined genomic intervals. The resulting VCF is subsequently decomposed into biallelic records, deduplicated, harmonized to the build-specific reference-site file, and filtered to remove records without a valid alternate allele or records in which ALT is identical to REF. Variant identifiers are then standardized using the same CHROM_POS_REF_ALT format.

Separate PRS score files are used for GRCh37 and GRCh38. Each score file contains a variant identifier, an effect allele, and score-weight columns. During scoring, PLINK2 [4] matches variants in the harmonized WGS genotype data to the effect allele specified in the corresponding score file and applies the associated weights to calculate each PRS.

### PRS quality control

Before PRS calculation, quality control is performed by comparing variant identifiers in the processed VCF with those required by the build-specific score files. PLINK2 --write-snplist is used to obtain the set of unique variants available in the processed VCF. For each score file, the workflow records the total number of unique score variants, the number absent from the VCF, and the corresponding missing-variant percentage. The same statistics are calculated for the union of variants across both score files.

Variant availability is additionally evaluated separately for each PRS. For each score, the pipeline determines the total number of variants in the PRS definition $N_{total}$, the number absent from the individual's processed VCF $N_{missing}$, and the number available for score calculation:

$$N_{used} = N_{total} - N_{missing}$$

The resulting $N_{used}$ values are retained as PRS-specific denominators for subsequent score normalization. The application reports individual-PRS-level missingness, allowing the completeness of each calculated PRS to be assessed. Scores with more than 25% of missing variants are removed.

### PRS calculation

PRSs are calculated using PLINK2 --score, with score sums retained (cols=+scoresums,-scoreavgs). All score columns following the variant-identifier and effect-allele columns are processed, allowing multiple PRSs to be calculated in parallel from the same score file.

For each PRS, the combined PLINK2 score sum is normalized according to the number of score variants available in the processed VCF:

$$PRS_{norm} = \frac{PRS_{raw}}{2 * N_{used}}$$

Where $PRS_{raw}$ is the combined PLINK2 weighted allele-dosage sum for PRS, and. This normalization accounts for differences in score-variant availability across individual genotype files.

### Population structure adjustment

To ensure consistency between PRSs calculated from individual whole-genome sequencing data and those used for disease-model development in UK Biobank, PrognosIQ applies the same population-structure adjustment procedure used during model training. Principal-component scores are obtained by projecting the input genotype data onto the build-specific PCA reference axes derived from the 1000 Genomes Project using HapMap3 variants. The first six projected principal components (PC1-PC6) are used for ancestry adjustment.

For each PRS, PrognosIQ applies a pre-fitted linear regression model derived in UK Biobank that predicts the normalized PRS from PC1-PC6. The PC-associated component predicted by this model is subtracted from the normalized PRS:

$$PRS_{adj} = PRS_{norm} - PRS_{predicted\ (PC1-PC6)}$$

Thus, $PRS_{predicted(PC1\text{-}PC6)}$ is the PRS value predicted from the individual's first six projected principal components using the corresponding UK Biobank-derived regression model. The resulting $PRS_{adj}$ therefore represents the component of the normalized PRS remaining after adjustment for population structure.

The population-adjusted PRS is subsequently standardized using the corresponding UK Biobank reference mean and standard deviation:

$$PRS_{standardized} = \frac{PRS_{adj} - \mu_{UK\ Biobank}}{\sigma_{UK\ Biobank}}$$

where $\mu_{\text{UK Biobank}}$ and $\sigma_{\text{UK Biobank}}$ are the reference mean and standard deviation for the corresponding population-adjusted PRS derived from UK Biobank. The resulting standardized PRS is used as the genetic predictor in the corresponding PrognosIQ disease-risk model.

#### Calculation of PRS in Biobank Russia

PRSs in Biobank Russia [10] were calculated using the same general scoring and preprocessing framework as that applied in UK Biobank. Individual PRSs were normalized according to the number of variants contributing to each score. Population structure was accounted for using a PCA-based residualization procedure analogous to that used in UK Biobank. Specifically, for each PRS, the normalized score was regressed on the first six projected genetic principal components (PC1-PC6) within Biobank Russia, and the fitted PC-associated component was subtracted from the normalized score. To ensure compatibility with the genetic feature scale used during development of the PrognosIQ prediction models, the resulting population-structure-adjusted PRSs were subsequently z-standardized using the corresponding reference means and standard deviations derived from UK Biobank. These standardized PRSs were then used as inputs to the PrognosIQ models for external validation in Biobank Russia.

#### PrognosIQ validation in Biobank Russia

Baseline Biobank Russia data were harmonized to the predictor definitions used by the corresponding PrognosIQ disease models. Available demographic, anthropometric, laboratory, smoking, and medical-history variables were mapped to the corresponding model features. For predictors required by a PrognosIQ model but not directly represented in Biobank Russia, corresponding variables were derived from the available phenotypic data where possible. Missing values in numeric predictors were imputed using the corresponding mean observed in Biobank Russia. Population-structure-adjusted PRSs were incorporated as genetic predictors as described above (**Sup. materials. Calculation of PRS in Biobank Russia**).

For type 2 diabetes, 10-year PrognosIQ risk predictions were generated from baseline clinical and genetic data using the corresponding clinical-genetic type 2 diabetes prediction model. Participants with history of type 2 diabetes or use of glucose-lowering medication at baseline were excluded from the prospective analysis. Incident type 2 diabetes during follow-up was defined using the available follow-up diagnosis variable.

PrognosIQ predictions were compared with FINDRISC calculated from the baseline variables available in Biobank Russia.

For ASCVD, PrognosIQ predictions were generated separately for myocardial infarction, major coronary heart disease events, and stroke using the corresponding disease-specific models. The disease-specific 10-year predictions were combined to obtain an overall PrognosIQ ASCVD prediction for each participant. Participants with cardiovascular disease or statin use at baseline were excluded from the prospective ASCVD analysis. Incident ASCVD was defined as a composite outcome comprising myocardial infarction, stroke or transient ischemic attack, coronary revascularization, or cardiovascular death. A participant was classified as an ASCVD case if any component of the composite outcome occurred during follow-up.

For comparison, 10-year PREVENT-ASCVD risk was calculated using the PooledCohort R package package (v0.0.2) [11]. Inputs included age, sex, current smoking status, total and high-density lipoprotein cholesterol, systolic blood pressure, antihypertensive and statin medication use, diabetes status, body mass index, and estimated glomerular filtration rate. PREVENT-ASCVD eligibility and input-variable ranges were applied before calculation of the score.

Within each disease-specific analysis, PrognosIQ and the corresponding established clinical risk score were evaluated in the same participants. Discrimination for incident disease over the 10-year follow-up period was assessed using ROC AUC, calculated in R using the pROC package (v1.18.5) [12].

### Supplementary figures

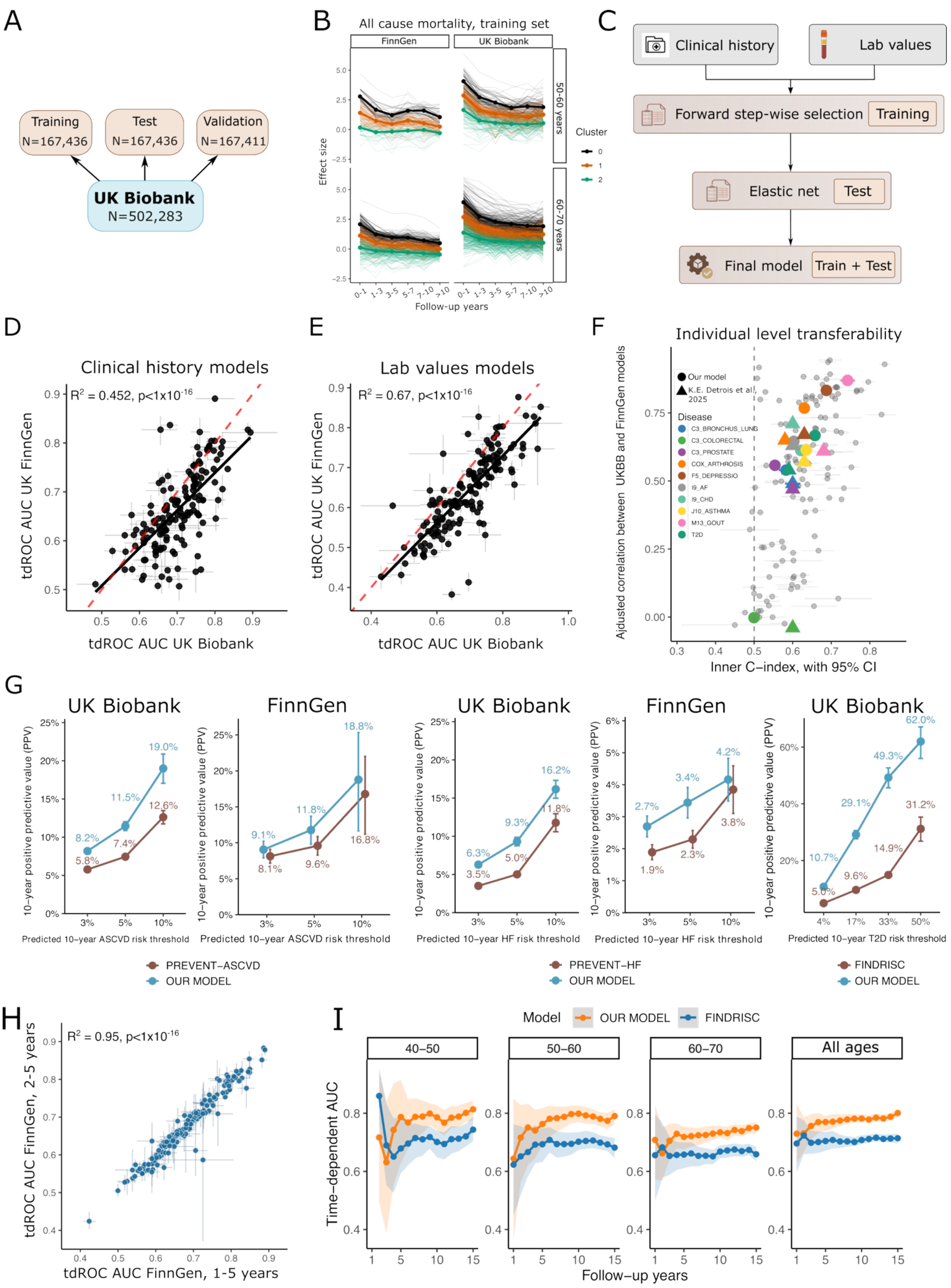
A
Training N=167,436
Test N=167,436
Validation N=167,411
UK Biobank N=502,283
B
All cause mortality, training set
FinnGen
UK Biobank
50-60 years
60-70 years
Effect size
Cluster
0
1
2
Follow-up years
0-1
1-3
3-5
5-7
7-10
>10
C
Clinical history
Lab values
Forward step-wise selection
Training
Elastic net
Test
Final model
Train + Test
D
Clinical history models
R² = 0.452, p<1x10⁻¹⁶
tdROC AUC UK FinnGen
tdROC AUC UK Biobank
E
Lab values models
R² = 0.67, p<1x10⁻¹⁶
tdROC AUC UK FinnGen
tdROC AUC UK Biobank
F
Individual level transferability
Our model
K.E. Detrois et al., 2025
Disease
C3_BRONCHUS_LUNG
C3_COLORECTAL
C3_PROSTATE
COX_ARTHROSIS
F5_DEPRESSIO
I9_AF
I9_CHD
J10_ASTHMA
M13_GOUT
T2D
Ajdusted correlation between UKBB and FinnGen models
Inner C−index, with 95% CI
G
UK Biobank
FinnGen
UK Biobank
FinnGen
UK Biobank
10−year positive predictive value (PPV)
Predicted 10−year ASCVD risk threshold
Predicted 10−year ASCVD risk threshold
Predicted 10−year HF risk threshold
Predicted 10−year HF risk threshold
Predicted 10−year T2D risk threshold
PREVENT−ASCVD
OUR MODEL
PREVENT−HF
OUR MODEL
FINDRISC
OUR MODEL
H
R² = 0.95, p<1x10⁻¹⁶
tdROC AUC FinnGen, 2-5 years
tdROC AUC FinnGen, 1-5 years
I
Model
OUR MODEL
FINDRISC
40−50
50−60
60−70
All ages
Time−dependent AUC
Follow−up years

**Supplementary Figure S1. Development of disease-specific clinical prediction models.**

**A) Partitioning of the UK Biobank cohort.** The UK Biobank cohort (N = 502,283) was divided into training (N = 167,436), test (N = 167,436), and validation (N = 167,411) datasets. The training and test datasets were used for model development and optimization, whereas the validation dataset was reserved for evaluating predictive performance.

**B) Temporal patterns of associations between clinical-history phenotypes and all-cause mortality.** Associations were estimated in the training datasets using Cox models with time-dependent exposures and coefficients varying across time-since-diagnosis intervals: 0-1, 1-3, 3-5, 5-7, 7-10, and >10 years. Panels show FinnGen and UK Biobank separately for the 50-60- and 60-70-year baseline-age groups. Thin lines represent the estimated association profiles of individual clinical-history phenotypes, expressed as Cox regression coefficients on the log-hazard scale. Phenotypes were grouped into three clusters according to their temporal profiles; thick lines with points indicate the cluster-specific mean profiles, colored black, orange, and green.

**C) Clinical model-development workflow.** Candidate predictors comprised clinical-history phenotypes and routine laboratory measurements. Forward stepwise feature selection was performed in the training dataset, followed by elastic-net regularization, with regularization parameters selected according to predictive performance in the test dataset. Selected predictors were used to fit the final log-normal accelerated failure-time model in the combined training and test datasets.

**D) Cross-biobank transferability of clinical-history model performance.** Each point represents a disease-specific model evaluated in the UK Biobank and FinnGen validation datasets. The x-axis shows mean time-dependent receiver operating characteristic area under the curve (tdROC AUC) across prediction horizons of 1-5 years in UK Biobank, and the y-axis shows the corresponding estimate in FinnGen. Horizontal and vertical error bars indicate 95% percentile bootstrap confidence intervals based on 100 bootstrap resamples. The solid black line represents the fitted linear regression across disease-specific performance estimates, and the dashed red diagonal indicates equal performance in the two cohorts. The coefficient of determination ($R^2$) and corresponding P value summarize the association between performance estimates across biobanks.

**E) Cross-biobank transferability of laboratory-value model performance.** Disease-specific models based on laboratory measurements were evaluated using the same performance measures and graphical conventions as in panel D.

**F) Individual-level transferability of disease-specific prediction models.** The x-axis shows within-cohort discrimination, measured by the C-index, with horizontal error bars indicating 95% confidence intervals. The y-axis shows the correlation between individual-level scores generated by models trained independently in UK Biobank and FinnGen, after removing the model-specific contributions of baseline age and sex. Circles represent models developed in the present study, and triangles represent the published results of Detrois et al. [13]. Colors identify selected disease outcomes, as indicated in the legend; the remaining outcomes are shown in gray. The vertical dashed line marks a C-index of 0.5.

**G) Positive predictive value at selected risk thresholds.** Panels compare the developed clinical models with PREVENT-ASCVD for atherosclerotic cardiovascular disease (ASCVD) in UK Biobank and FinnGen, PREVENT-HF for heart failure in UK Biobank and FinnGen, and FINDRISC for type 2 diabetes in UK Biobank, from left to right. Positive predictive value (PPV) was estimated as the Kaplan-Meier-derived 10-year cumulative incidence among participants classified at or above each risk threshold. ASCVD and heart failure comparisons use thresholds of 3%, 5%, and 10%; the type 2 diabetes comparison uses thresholds of 4%, 17%, 33%, and 50%. Blue points and lines represent the developed clinical models, and brown points and lines represent the established risk scores. Error bars indicate 95% confidence intervals.

**H) Sensitivity analysis excluding first-year incident cases in FinnGen.** Each point represents a disease outcome. The x-axis shows mean tdROC AUC across prediction horizons of 1-5 years in the primary analysis, and the y-axis shows mean tdROC AUC across horizons of 2-5 years after excluding participants diagnosed with the target disease within the first year after baseline. Horizontal and vertical error bars indicate 95% bootstrap confidence intervals. The dashed diagonal indicates equal performance estimates. $R^2$ denotes the squared Pearson correlation coefficient, and the P value was obtained from a two-sided Pearson correlation test.

**I) Prediction of type 2 diabetes among metabolically healthy UK Biobank participants.** Analyses were restricted to participants in the validation dataset without prevalent type 2 diabetes who had baseline glucose <5.5 mmol/L, HbA1c <42 mmol/mol, body mass index <30 kg/m², and no use of diabetes-related medications. Panels show participants aged 40-50, 50-60, and 60-70 years at baseline and the combined 40-70-year sample. The x-axis shows the prediction horizon in years, and the y-axis shows tdROC AUC. Orange curves represent the developed clinical model, and blue curves represent FINDRISC. Points indicate annual performance estimates, and shaded areas indicate 95% percentile bootstrap confidence intervals based on 100 bootstrap resamples.

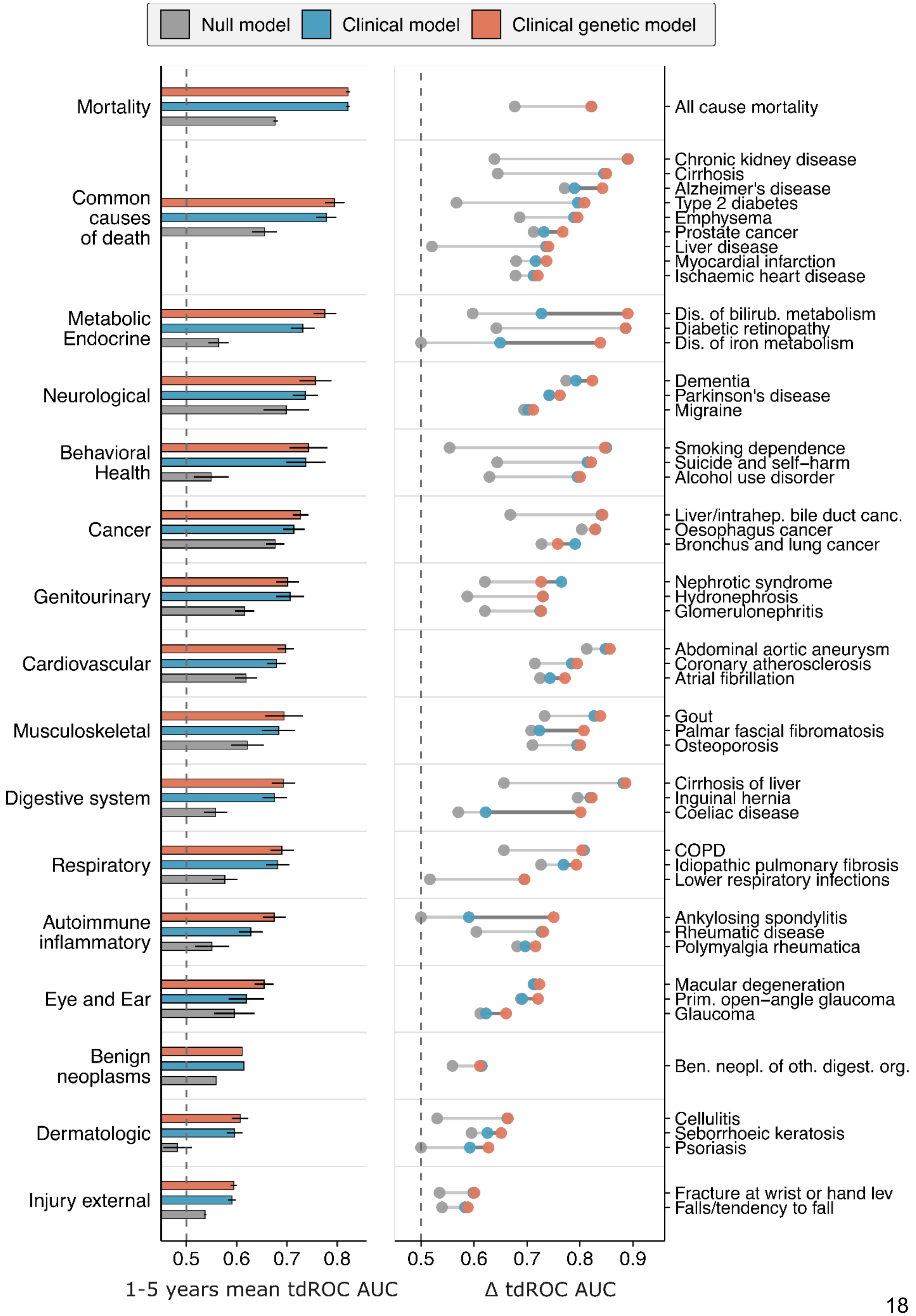

Null model
Clinical model
Clinical genetic model
Mortality
Common causes of death
Metabolic Endocrine
Neurological
Behavioral Health
Cancer
Genitourinary
Cardiovascular
Musculoskeletal
Digestive system
Respiratory
Autoimmune inflammatory
Eye and Ear
Benign neoplasms
Dermatologic
Injury external
All cause mortality
Chronic kidney disease
Cirrhosis
Alzheimer's disease
Type 2 diabetes
Emphysema
Prostate cancer
Liver disease
Myocardial infarction
Ischaemic heart disease
Dis. of bilirub. metabolism
Diabetic retinopathy
Dis. of iron metabolism
Dementia
Parkinson's disease
Migraine
Smoking dependence
Suicide and self−harm
Alcohol use disorder
Liver/intrahep. bile duct canc.
Oesophagus cancer
Bronchus and lung cancer
Nephrotic syndrome
Hydronephrosis
Glomerulonephritis
Abdominal aortic aneurysm
Coronary atherosclerosis
Atrial fibrillation
Gout
Palmar fascial fibromatosis
Osteoporosis
Cirrhosis of liver
Inguinal hernia
Coeliac disease
COPD
Idiopathic pulmonary fibrosis
Lower respiratory infections
Ankylosing spondylitis
Rheumatic disease
Polymyalgia rheumatica
Macular degeneration
Prim. open−angle glaucoma
Glaucoma
Ben. neopl. of oth. digest. org.
Cellulitis
Seborrhoeic keratosis
Psoriasis
Fracture at wrist or hand lev
Falls/tendency to fall
0.5
0.6
0.7
0.8
0.9
1-5 years mean tdROC AUC
Δ tdROC AUC

**Supplementary Figure S2. Predictive performance of the null, clinical, and clinical-genetic models in the FinnGen external validation cohort.** Each model's performance was calculated as the mean 1-5-year tdROC AUC. The left panel shows mean model performance across disease categories, whereas the right panel shows performance for representative individual diseases within each category. For disease-category summaries, error bars indicate the standard error of the mean tdROC AUC across diseases within each category; for all-cause mortality, the disease-specific confidence interval is shown. Diseases were displayed if either the clinical or clinical-genetic model showed a significant positive improvement over the null model after FDR correction, and up to 3 diseases with the highest clinical-genetic model performance were displayed per category; all prespecified common causes of death were retained.

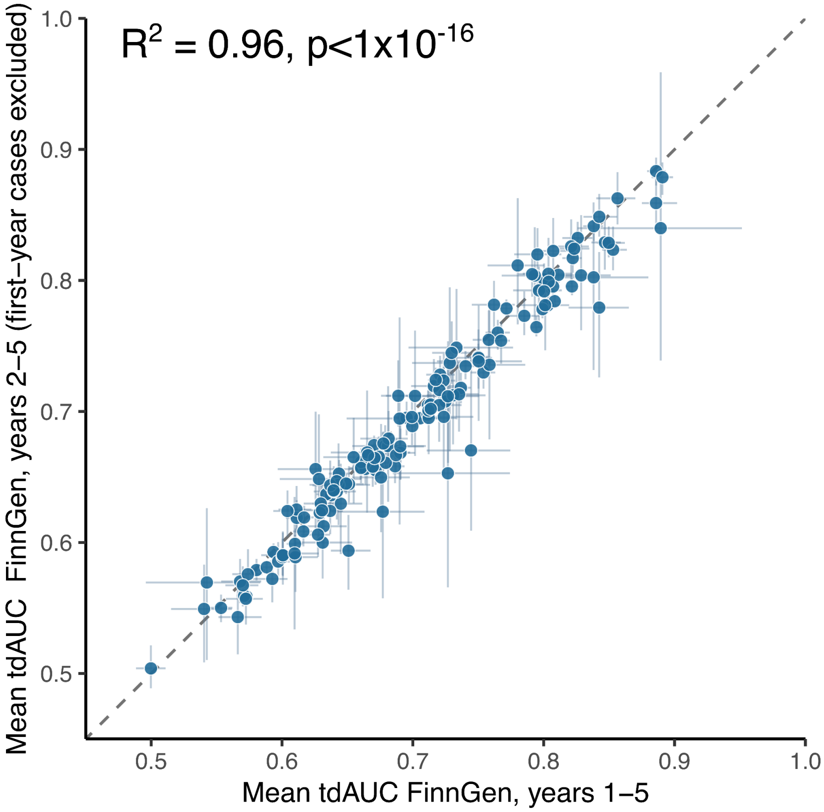


**Supplementary Figure S3. Sensitivity analysis of clinical-genetic model performance after excluding first-year incident cases in FinnGen.** Each point represents an outcome evaluated among participants aged 40-70 years. The x-axis shows mean 1-5-year time-dependent ROC AUC (tdROC AUC) in the primary analysis, and the y-axis shows mean 2-5-year tdROC AUC after excluding participants who experienced the target outcome within the first year after baseline. Horizontal and vertical error bars indicate 95% percentile bootstrap confidence intervals. The dashed diagonal represents equal predictive performance. $R^2$ denotes the squared Pearson correlation coefficient between performance estimates across outcomes; the P value was obtained from a two-sided Pearson correlation test.

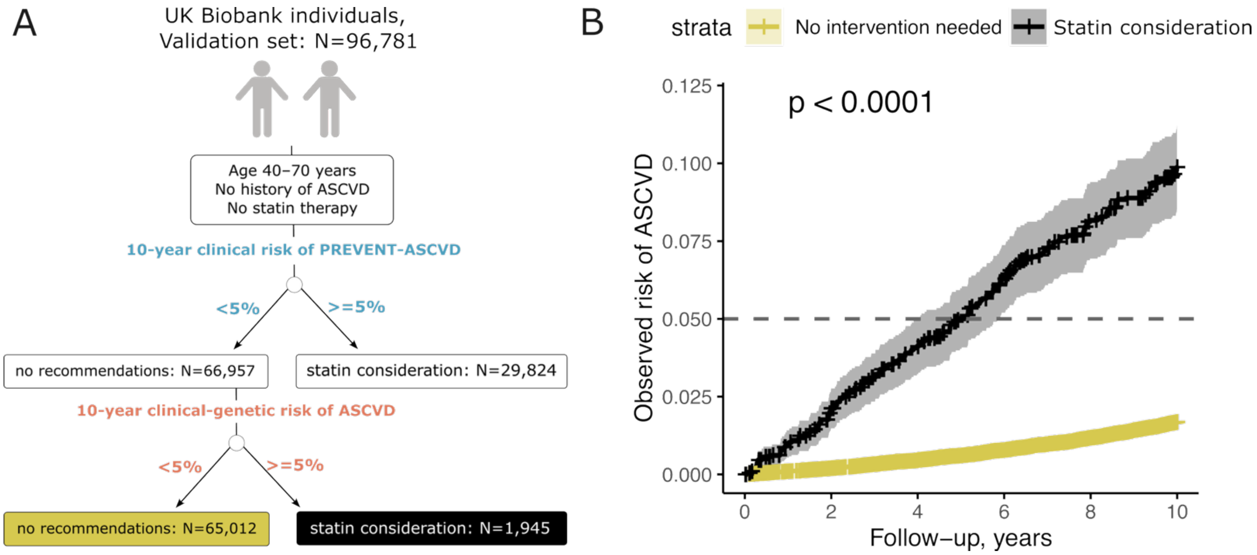


**Supplementary Figure S4. Clinical-genetic risk reclassification across the 5% PREVENT-ASCVD risk threshold identifies groups with distinct observed risk in UK Biobank.**

**A)** Reclassification of 10-year PREVENT-ASCVD risk after incorporation of genetic information in 96,781 UK Biobank participants aged 40-70 years with no history of ASCVD and no prior statin therapy. Based on the clinical model, 66,957 participants had a predicted 10-year ASCVD risk <5% and 29,824 had a risk ≥5%. After incorporation of genetic information, 1,945 participants were reclassified upward from <5% to ≥5%

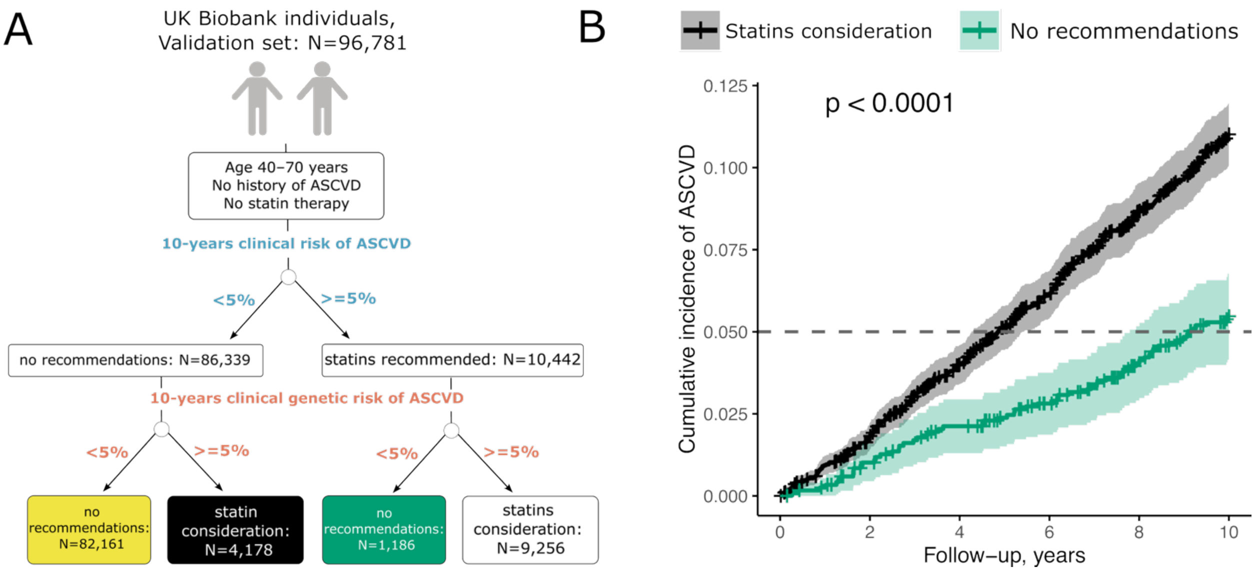


**Supplementary Figure S5. Clinical-genetic risk reclassification across the 5% ASCVD risk threshold identifies groups with distinct observed risk in UK Biobank.**

**A)** Reclassification of 10-year ASCVD risk after incorporation of genetic information in 96,781 UK Biobank participants aged 40-70 years with no history of ASCVD and no prior statin therapy. Based on the clinical model, 86,339 participants had a predicted 10-year ASCVD risk <5% and 10,442 had a risk ≥5%. After incorporation of genetic information, 4,178 participants were reclassified upward from <5% to ≥5%, whereas 1,186 participants were reclassified downward from ≥5% to <5%. The remaining 82,161 and 9,256 participants remained below and above the 5% threshold, respectively.

**B)** Kaplan-Meier estimates of cumulative ASCVD incidence among the two discordantly reclassified groups. Participants reclassified upward from a clinical risk <5% to a clinical-genetic risk ≥5% had substantially higher observed ASCVD risk than participants reclassified downward from a clinical risk ≥5% to a clinical-genetic risk <5%. Shaded areas indicate 95% confidence intervals. The horizontal dashed line indicates the 5% ASCVD risk threshold.

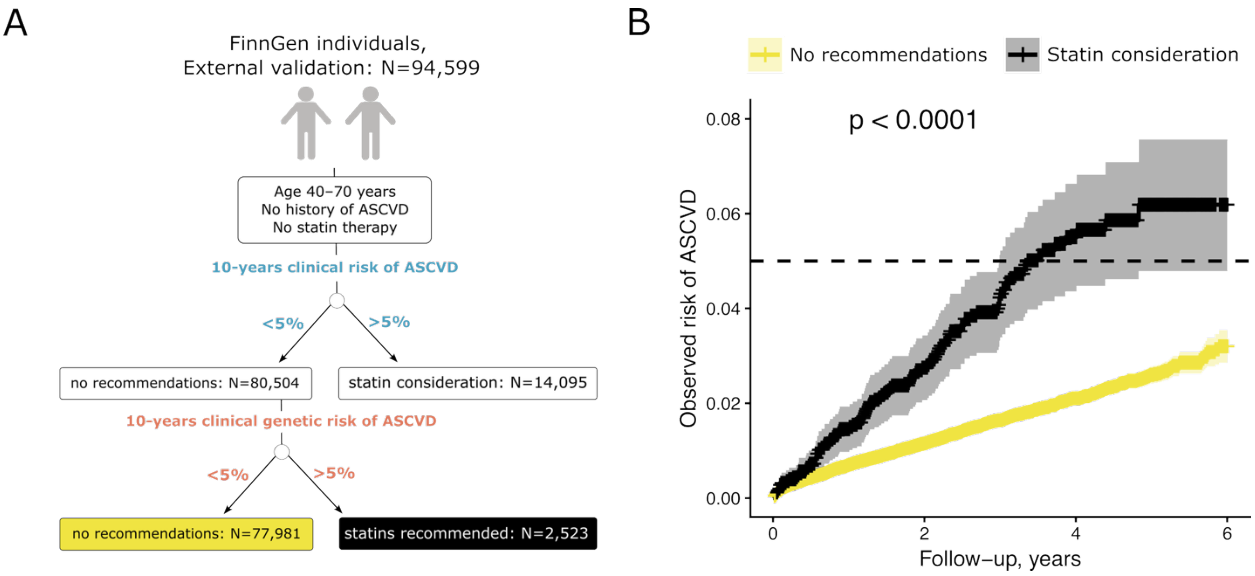


**Supplementary Figure S6. Clinical-genetic risk reclassification across the 5% ASCVD risk threshold identifies groups with distinct observed risk in FinnGen.**

**A)** Reclassification of 10-year ASCVD risk after incorporation of genetic information in 94,599 FinnGen participants aged 40-70 years with no history of ASCVD and no prior statin therapy. Based on the clinical model, 80,504 participants had a predicted 10-year ASCVD risk <5% and 14,095 had a risk ≥5%. After incorporation of genetic information, 2,523 participants were reclassified upward from <5% to ≥5%

**B)** Kaplan-Meier estimates of cumulative ASCVD incidence among the two discordantly reclassified groups. Participants reclassified upward from a clinical risk <5% to a clinical-genetic risk ≥5% had substantially higher observed ASCVD risk than participants without reclassification. Shaded areas indicate 95% confidence intervals. The horizontal dashed line indicates the 5% ASCVD risk threshold.

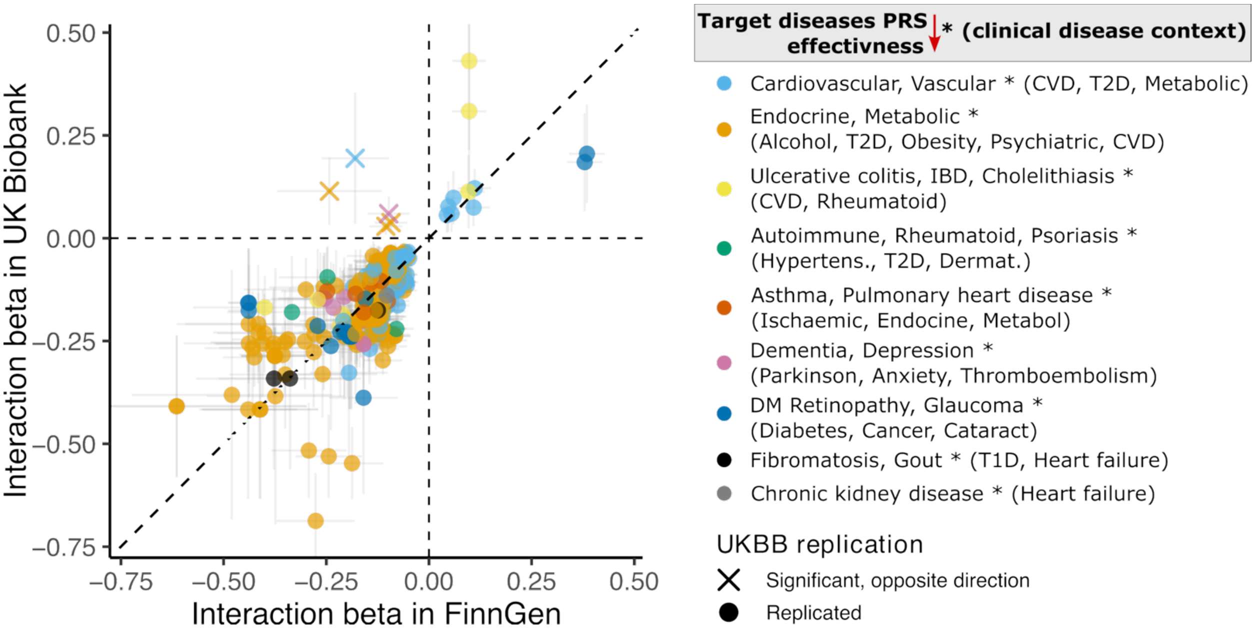


**Supplementary Figure S7.** Replication of clinic-genetic interaction effects across biobanks. Horizontal and vertical error bars indicate 95% confidence intervals for the interaction effect estimates in FinnGen and UK Biobank, respectively. The dashed diagonal line indicates equal interaction effect sizes in the two cohorts.

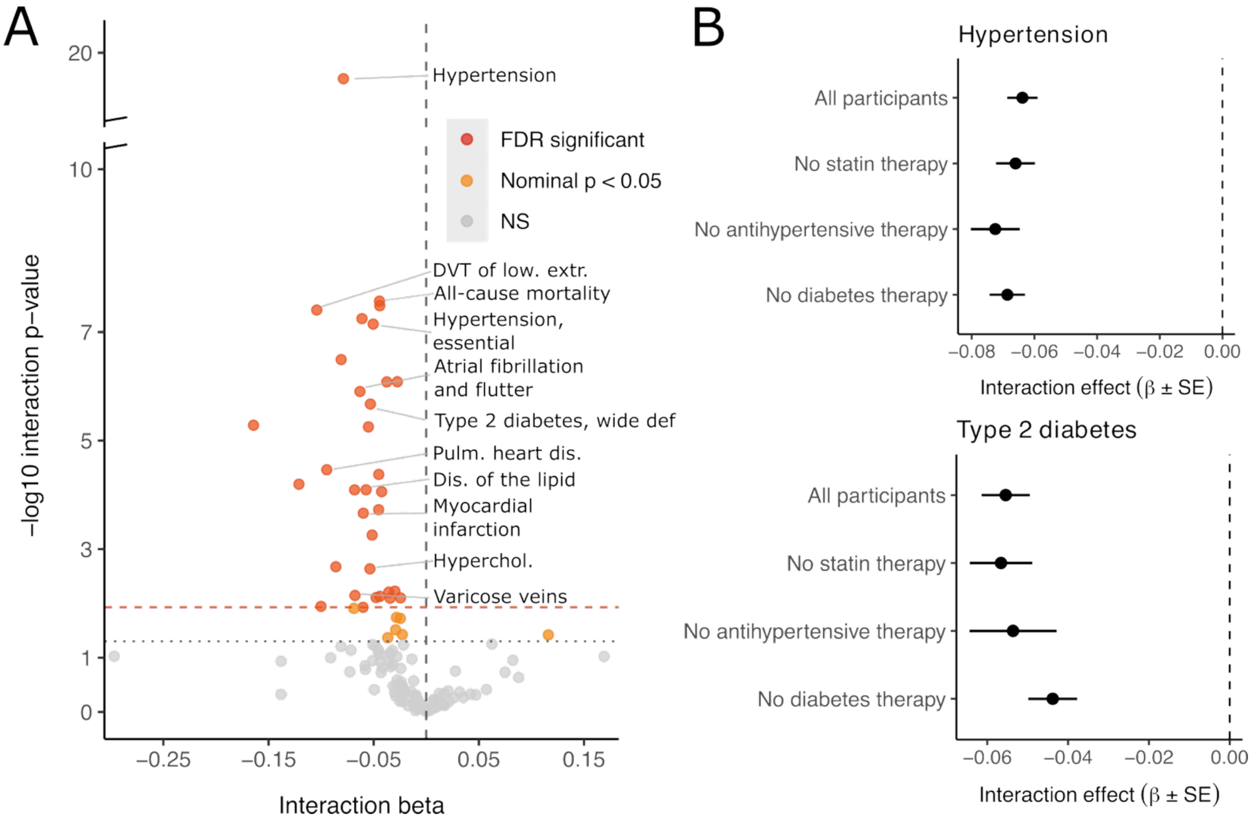

**Supplementary Figure S8. Interaction between polygenic risk and clinical risk, with sensitivity analyses excluding medication users.**
**A)** PRS–clinical risk interaction effects across phenotypes. The x-axis shows the interaction coefficient (β) between PRS and the clinical linear predictor, and the y-axis shows the −log10 interaction P value. The vertical dashed line denotes no interaction (β = 0), while horizontal reference lines indicate nominal and FDR-adjusted significance thresholds. Selected phenotypes with strong or significant interaction effects are labeled.
**B)** Sensitivity analyses for hypertension (top) and type 2 diabetes (bottom) show interaction coefficients estimated in all participants and after excluding individuals receiving statin, antihypertensive, or diabetes therapy. Points represent interaction β estimates and horizontal bars indicate ±1 standard error; the vertical dashed line denotes β = 0.

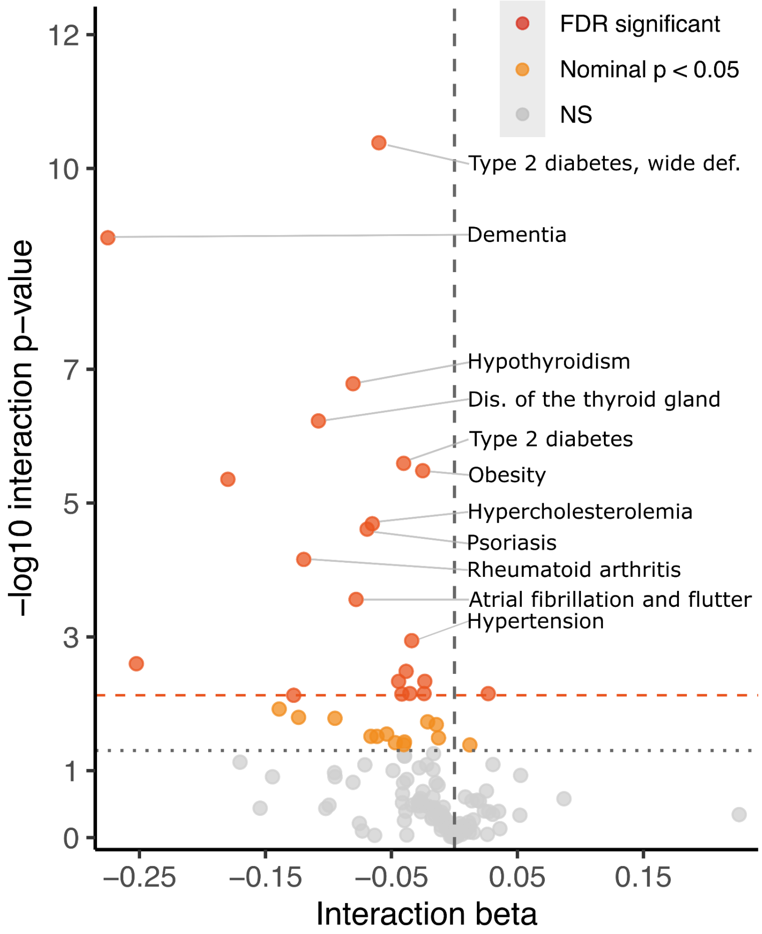


**Supplementary Figure S9. Interaction between polygenic risk and clinical risk after accounting for PRS-by-age and PRS-by-sex interactions.**
**A)** PRS-clinical risk interaction effects across phenotypes. The x-axis shows the interaction coefficient (β) between PRS and the clinical linear predictor, and the y-axis shows the −log10 interaction P value. The vertical dashed line denotes no interaction (β

= 0), while horizontal reference lines indicate nominal and FDR-adjusted significance thresholds. Selected phenotypes with strong or significant interaction effects are labeled.

### References


1. Bycroft C, Freeman C, Petkova D, Band G, Elliott LT, Sharp K, Motyer A, Vukcevic D, Delaneau O, O'Connell J, Cortes A, Welsh S, Young A, Effingham M, McVean G, Leslie S, Allen N, Donnelly P, Marchini J. The UK Biobank resource with deep phenotyping and genomic data. Nature. 2018 Oct;562(7726):203-209. doi: 10.1038/s41586-018-0579-z. Epub 2018 Oct 10. PMID: 30305743; PMCID: PMC6786975.
2. Lambert SA, Gil L, Jupp S, et al. The Polygenic Score Catalog as an open database for reproducibility and systematic evaluation. Nature Genetics. 2021;53:420–425. doi:10.1038/s41588-021-00783-5.
3. Lambert SA, Wingfield B, Gibson JT, et al. Enhancing the Polygenic Score Catalog with tools for score calculation and ancestry normalization. Nature Genetics. 2024;56:1989–1994. doi:10.1038/s41588-024-01937-x
4. Chang CC, Chow CC, Tellier LC, Vattikuti S, Purcell SM, Lee JJ. Second-generation PLINK: rising to the challenge of larger and richer datasets. GigaScience. 2015;4:7. doi:10.1186/s13742-015-0047-8.
5. The 1000 Genomes Project Consortium. A global reference for human genetic variation. Nature. 2015;526:68–74. doi:10.1038/nature15393
6. Hao L, Kraft P, Berriz GF, et al. Development of a clinical polygenic risk score assay and reporting workflow. Nature Medicine. 2022;28:1006–1013.
7. Gerds TA. (2025). pec: Prediction Error Curves for Risk Prediction Models in Survival Analysis. R package version 2025.06.24. doi:10.32614/CRAN.package.pec.
8. Danecek P, Bonfield JK, Liddle J, et al. Twelve years of SAMtools and BCFtools. GigaScience. 2021;10(2):giab008. doi:10.1093/gigascience/giab008.
9. Van der Auwera GA, O'Connor BD. Genomics in the Cloud: Using Docker, GATK, and WDL in Terra. 1st ed. O'Reilly Media; 2020.
10. Usoltsev, D., Kolosov, N., Rotar, O. et al. Complex trait susceptibilities and population diversity in a sample of 4,145 Russians. Nat Commun 15, 6212 (2024). https://doi.org/10.1038/s41467-024-50304-1
11. Jaeger, B. C. PooledCohort: Predicted Risk for CVD Using Pooled Cohort Equations, PREVENT Equations, and Other Contemporary CVD Risk Calculators. R package version 0.0.2 (2024). https://CRAN.R-project.org/package=PooledCohort

12. Robin X, Turck N, Hainard A, Tiberti N, Lisacek F, Sanchez JC, Müller M. pROC: an open-source package for R and S+ to analyze and compare ROC curves. BMC Bioinformatics. 2011;12:77. doi:10.1186/1471-2105-12-77
13. Detrois, K.E., Hartonen, T., Teder-Laving, M. et al. Cross-biobank generalizability and accuracy of electronic health record-based predictors compared to polygenic scores. Nat Genet 57, 2136–2145 (2025). https://doi.org/10.1038/s41588-025-02298-9
